# Neural Correlates of Human Food Memory link to Microbial, Homeostatic, and Hedonic Signals: Evidence from a Prebiotic Randomized Clinical Trial

**DOI:** 10.64898/2026.06.14.26355607

**Authors:** Daria E. A. Jensen, Ronja Thieleking, Evelyn Medawar, Madlen Reinicke, Ulrike Rolle-Kampczyk, Martin von Bergen, Michael Stumvoll, Arno Villringer, Frauke Beyer, A. Veronica Witte

## Abstract

**Background:** Homeostatic and hedonic brain circuits regulate eating behavior but also shape how food memories are encoded and retrieved.

**Objective:** We examined neural correlates during food memory encoding and retrieval during functional MRI before and after a 14-day prebiotic intervention in a preregistered, double-blind crossover trial (NCT03829189).

**Design:** 55 healthy adults with overweight (19 females, age 28±6.5, BMI 25-30 kg/m^2^) underwent 3 Tesla task-based functional MRI before and after dietary intervention of prebiotic (30g inulin/day) or equicaloric placebo for 14 days. Peripheral metabolic, short-chain fatty acids (SCFA), and microbial markers using 16S rRNA analysis were assessed in fasting blood and feces.

**Results:** Food memory was enhanced by assigned reward value and engaged brain activity in hedonic regions, including the nucleus accumbens, orbitofrontal cortex, caudate, cingulate, dorsomedial prefrontal cortex, and ventral tegmental area, as well as homeostatic and memory-related such as the hypothalamus and the hippocampus. Higher neural activations during food encoding were related to higher *Actinobacteriota* abundance, fecal SCFA acetate, and creatinine levels, and lower ghrelin levels. Activations in reward-related and homeostatic brain areas partially correlated with insulin, glucagon-like peptide-1, leptin, and thyroid-stimulating hormone levels. Neural activations related to food memory decreased after prebiotic intervention. The prebiotic supplementation induced decrease of hippocampal activity during food encoding related to changes in gut microbiota *Firmicutes* abundance.

**Conclusions:** This study indicates that neuronal food-related memory processes depend on homeostatic and hedonic brain signals modulated by the gut-brain axis. Our findings raise implications for the treatment of obesity and substance use disorder.

## Introduction

Memory function is pivotal in food decision-making, as memory deficits can lead to reduced recall of recent eating episodes, influencing subsequent meal decisions^1,2^. Additionally, positive and negative memories of past food encounters and environmental determinants are essential for developing food preferences^3^. Memorizing eating and the consequences of consuming certain foods, such as fullness, texture, taste or well-being, involve both homeostatic (energy-balance) and hedonic (reward-related) processes^4,5^. However, the underlying mechanisms and neural correlates of food memory, as opposed to non-food memory, remain to be fully understood.

The hippocampus has been consistently linked to memory performance^6,7^ and food intake control^8^. For instance, memory-related hippocampal activity measured with functional magnetic resonance imaging (fMRI) during food cue processing was more pronounced in women with overweight and obesity^9^, and lesion studies showed that hippocampal damage can lead to excessive food intake and weight gain.

The hypothalamus interacts with the hippocampus to encode food-related experiences^8^. It controls homeostatic regulation of food intake by processing signals from the gut, adipose tissue, and brainstem to maintain energy balance^5,10^. For example, via the hypothalamus, ghrelin (often called “hunger hormone”)^11^ was shown to enhance hippocampal function in mice^12^; while the gut peptide YY3-36 (PYY)^13^ and glucagon-like peptide-1 (GLP-1)^14^ signal satiety to terminate a meal. Insulin regulates glucose metabolism and stimulates the hypothalamus and prefrontal brain areas to reduce food intake^15^. Moreover, leptin (satiety hormone) is thought to accelerate long-term meal termination^16^. Leptin-deficient rodents exhibit impairments in hippocampal synaptic plasticity and spatial memory tasks^1–3^. Other metabolites such as creatinine, liver enzymes, and thyroid-stimulating hormone (TSH) are suggested to modulate hypothalamic^18,19^ and hippocampus activity^20–22^.

Notably, some of those homeostasis-related hormones and peptides have been linked to higher-order cognition (insulin^23,24^; GLP-1^25^; creatine^18^), yet their association with neural correlates of food memory in humans remains unclear. In addition, homeostatic and hedonic mechanisms are only partially distinct; for example, GLP-1 induces satiety through hypothalamic signals (“homeostatic”), but GLP-1 receptor agonists have also been related to hedonic inhibition, likely through dopaminergic signaling^26^.

Hedonic processes are linked to dopaminergic and serotonergic neurotransmission, and dopamine and 5-HT can boost the formation of memory^27,28^. These mesolimbic systems, include the ventral tegmental area (VTA), nucleus accumbens (NAc), and orbitofrontal cortex (OFC), and play a crucial role in encoding the reward value of food^29,30^. We have shown that memory performance for food stimuli improved with a higher reward value assigned to the respective stimuli^31^, and highly desirable foods activated reward-related brain regions^32^. However, experimental data on the specific role of hedonic processes on food memory and its neural correlates is scarce.

Emerging research suggests that also gut microbiota significantly influences food-related cognitive processes, including memory and reward sensitivity^33^. Gut-derived metabolites, such as short-chain fatty acids (SCFAs), modulate hippocampal function and dopamine signaling, potentially affecting both homeostatic and hedonic food memory^34^. Previous findings suggest that prebiotic supplementation with high-dose dietary fiber alters gut microbial diversity and fecal SCFA levels^35,36^, and modulates brain activation during food-related decision-making^32^.

In sum, the intricate relationship between memory function and food decision-making is likely influenced by both homeostatic and hedonic processes in the hypothalamus, reward-related brain areas, and the hippocampus. Additionally, previous research proposed a modulating role of the gut-brain axis, including gut microbiota and their metabolites in these processes. However, the specific neural pathways through which homeostatic and hedonic factors interact to influence food memory and decision-making remain unknown.

Thus, in this preregistered study (https://osf.io/whbc8), we aimed to identify the neural correlates of food memory during encoding and retrieval, using advanced task-based fMRI, and decipher the influence of peripheral metabolic and microbial markers on food memory-related brain activity patterns. We further investigated the effects of a prebiotic intervention on food-related memory in a randomized clinical trial (NCT03829189) of healthy adults with overweight. Our findings may help to develop novel intervention and management strategies for maladaptive eating behaviors, which are particularly relevant in today’s obesogenic environments^37^.

## Results

We examined food memory using a version of the mnemonic similarity task known to involve hippocampus function^38^ adapted to rewarding food and art stimuli^32,39^ at 3T MRI, in 55 healthy adults (19 females, 36 males), aged 19 to 45 years (mean age 28.4+-6.5), with a BMI of 27.3 kg/m²+-1.6 at first study visits (Supplementary Table 1). Briefly, after overnight fasting and a standardized small meal, participants were instructed to rate 80 food and 80 art images while undergoing fMRI (encoding phase). Ratings displayed the subjective value, i.e. the desire (or “wanting”) to eat the shown food, or to get the shown art image as high-quality print-out, respectively, on an 8-point Likert scale. After a break of 20 minutes, participants were asked to recognize old, similar, and new visual food and art targets during fMRI by responding either “old” for previously seen (old) images or “new” for previously unseen (similar, new) images (retrieval phase; see Figure 1A). All 55 participants performed above chance at food and art memory encoding (i.e., percentage of correctly responding with “old” compared to incorrectly responding with “new” to previously seen images at memory recognition 20 min later was higher than 50%), as well as food memory retrieval (i.e., percentage of correct identification of target (“old”) vs. lures and novel stimuli (“new”) during retrieval), however, at art memory retrieval, 17 participants performed lower than chance with a mean of 0.46+-0.02% correct art retrieval judgments across sessions (Figure 1B). Descriptive statistics for memory accuracy are reported in Supplementary Table 2, and an overview of preregistered hypotheses addressed in this study and statistical outcomes is shown in Supplementary Table 3.

**Figure 1:**
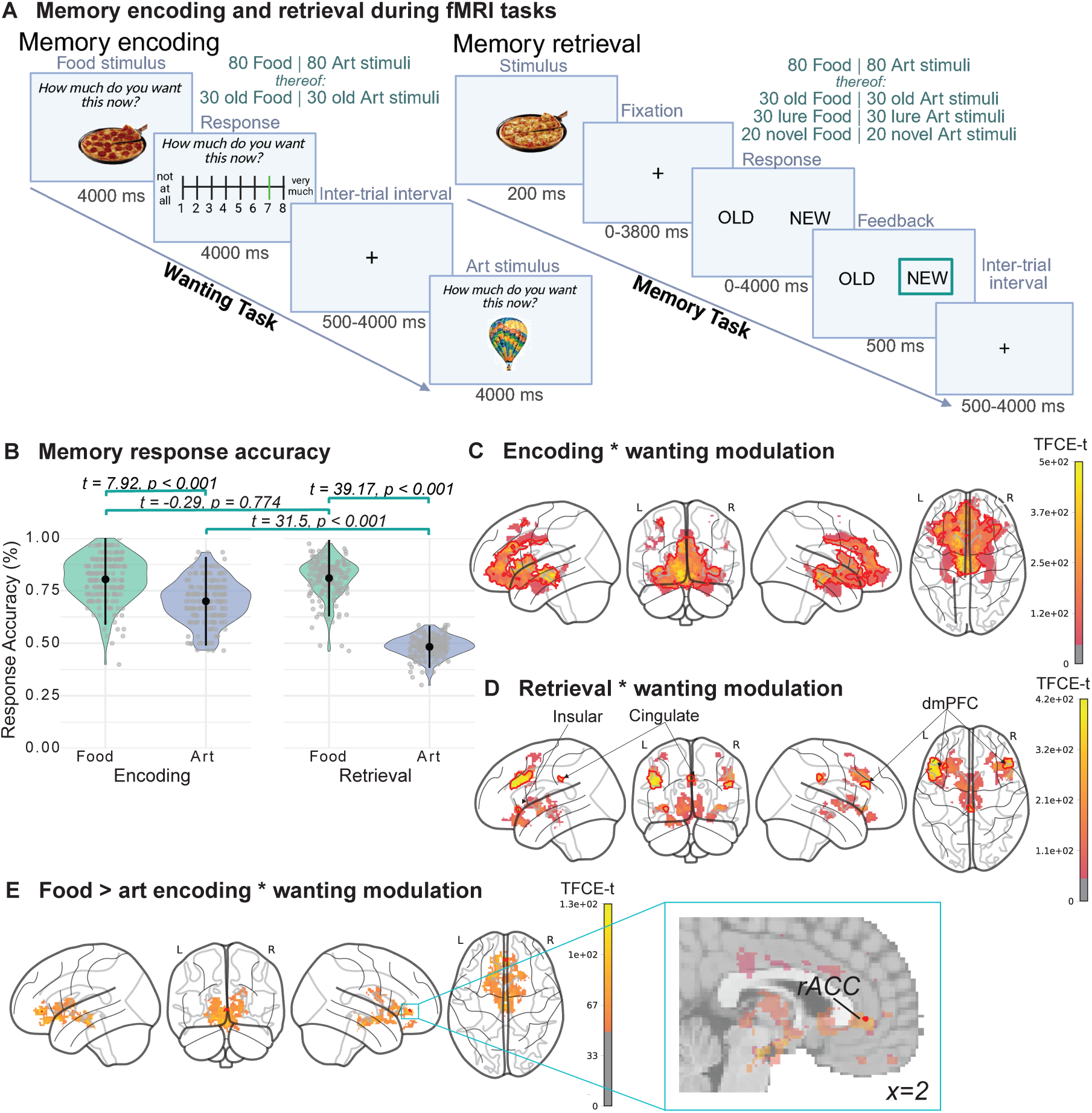
Response accuracy and neural response during memory encoding and retrieval of food and art stimuli. (A) Memory encoding during the ‘Wanting task’ and subsequent memory retrieval during the ‘Memory task’. Top: Memory encoding during the Wanting Task: Participants were asked to indicate on an 8-point Likert scale how much they want to have the depicted food or art image and to memorize the items. They were previously told that they would be rewarded with one of the highest-rated foods to eat and art to take away as a high-quality printout directly after the MRI scan. During this task, food and art images are encoded. Right: Memory retrieval during the memory task including target recognition and lure discrimination: Participants had to indicate as quickly and as accurately as possible if they had seen the presented food or art image in the previous task (“old”), if they had not seen it before (“new”) or if it was similar to a previously seen image (also “new”). Depicted are two exemplary, similar (but not identical) food and art stimuli. Originally created in BioRender.com. (B) Percentual response accuracy of art and food memory encoding and retrieval. We calculated the percentual response accuracy for food and art encoding (30 targets out of 80 per category and session) and retrieval (30 targets and 30+20 correct rejection stimuli per category and session). Out of 175 responses (55 participants x 4 sessions), one, four, three, and 47 subjects (100 entries for participant x sessions) performed below change level (<0.5) in food encoding, art encoding, food retrieval, and art retrieval, respectively. Differences between the mean response accuracy of food and art stimuli encoding and retrieval (top row) and between encoding and retrieval of art stimuli (second row, right) were estimated using t-tests and showed differences in their response accuracy (top row). However, no difference was shown between encoding and retrieval of food stimuli (second row, left). Differences in their mean were estimated using independent t-tests. Neural response during memory encoding and retrieval modulated by the wanting of individual food and art stimuli is shown in C-E. (C) Memory encoding and (D) memory retrieval (uncorrected for sex) were modulated by wanting scores and elicited in neural activations of large clusters during successful encoding and a small cluster during retrieval. (E) Food compared to art encoding was modulated by wanting scores and elicited neural activation in small clusters of the anterior cingulate cortex (ACC).

Successful encoding of both food and art stimuli was significantly associated with higher brain activations in hedonic and homeostatic brain areas, that is in the posterior OFC and NAc, and in the right thalamus and hypothalamus (*clusters 1-5, 8-3040 mm*^3^*, total: 3632 mm*^3^*; PTFCE-FWE<0.05-0.017*; Supplementary Figure 1A; Supplementary Table 4). During successful memory retrieval, we observed strong and widespread activations of over 50 cm^3^ extent, bilaterally in the OFC and NAc, and more frontal activations in the cingulate and paracingulate gyrus, all parts of the wider reward network and the hippocampus and amygdala (*cluster 1, 50128 mm*^3^*; PTFCE-FWE=0.001*; Supplementary Figure 1B). Successful versus non-successful memory retrieval of old images revealed specific bilateral activations in surrounding areas of the right and left caudate, posterior cingulate, and frontal pole (*clusters 1-3, 104-5400 mm*^3^*, total: 6136 mm*^3^*; PTFCE-FWE=0.002-0.042*; Supplementary Figure 1C). Further post-hoc analyses constrained to voxels in the hippocampus showed neural activations in the hippocampus head and hippocampus body during memory retrieval (*clusters 1-2, 872-2344 mm*^3^*, total: 3216 mm*^3^*; PTFCE-FWE=0.011-0.002*; Supplementary Figure 1D and Supplementary Table 5).

### Neural correlates of reward-enhanced memory

A higher wanting score (suggestive of subjective value) predicted higher activations in brain areas involved in both reward and homeostatic functions during memory encoding and retrieval (*H1_m* and *H1_e*, respectively; Table S4). Specifically, during encoding, large neural activations emerged for a range of reward areas, including the cingulate and dmPFC, but also the VTA, as well as homeostatic areas within the hypothalamus (Figure 1C). Those activations were slightly stronger in the left compared to the right hemisphere (*clusters 1-2, 104-38904 mm*^3^*, total: 39008 mm*^3^*; PTFCE-FWE=0.004-0.001*). Moreover, food compared to art reward-enhancement of memory encoding elicited neural activation of small clusters in the right anterior cingulate cortex (ACC; *clusters 1-2, 8-24 mm*^3^*, total: 32 mm*^3^*; PTFCE-FWE=0.049-0.022*; Figure 1E).

During retrieval, peak neural activations were identified in the left cingulate and insular cortex using voxel-wise inference (*clusters 1-2, 24-96 mm*^3^*, total: 120mm*^3^*; PFWE=0.043-0.012)*. These activations expanded to clusters in the dmPFC, VTA, and hypothalamus when applying a less stringent correction for covariates (without correction for sex and age, *clusters 1-4, 176-3056 mm*^3^*, total: 4168 mm*^3^*; PTFCE-FWE=0.02-0.009*; Figure 1D). We could not identify neural activations that were specifically related to food compared to art reward-enhancement of memory retrieval.

### Food-memory specific neural activations in reward areas and hippocampus

Next, we investigated food-specific memory activations independent of subjective wanting. When comparing neural activations for food versus art stimuli, we found that successful encoding elicited neural activations in a range of hedonic and reward-associated regions, including right pallidum, caudate, and NAc (*clusters 1-3, 88-8448 mm*^3^*, total: 8768 mm*^3^*; PTFCE-FWE =0.048-0.015*; Figure 2A, Supplementary Table 4, *H2_m*). Activations within the parahippocampus (cluster 6, 496 *mm*^3^*, PTFCE=0.001 of total: 42680 mm*^3^) and the hippocampus tail for food compared to art stimuli encoding using an explicit hippocampus mask (clusters 1-3, 32-320 mm^3^; total: 600mm3; *total 600mm*^3^*, PTFCE=0.0004; PTFCE-FWE>0.05*) did not survive FWE correction.

**Figure 2:**
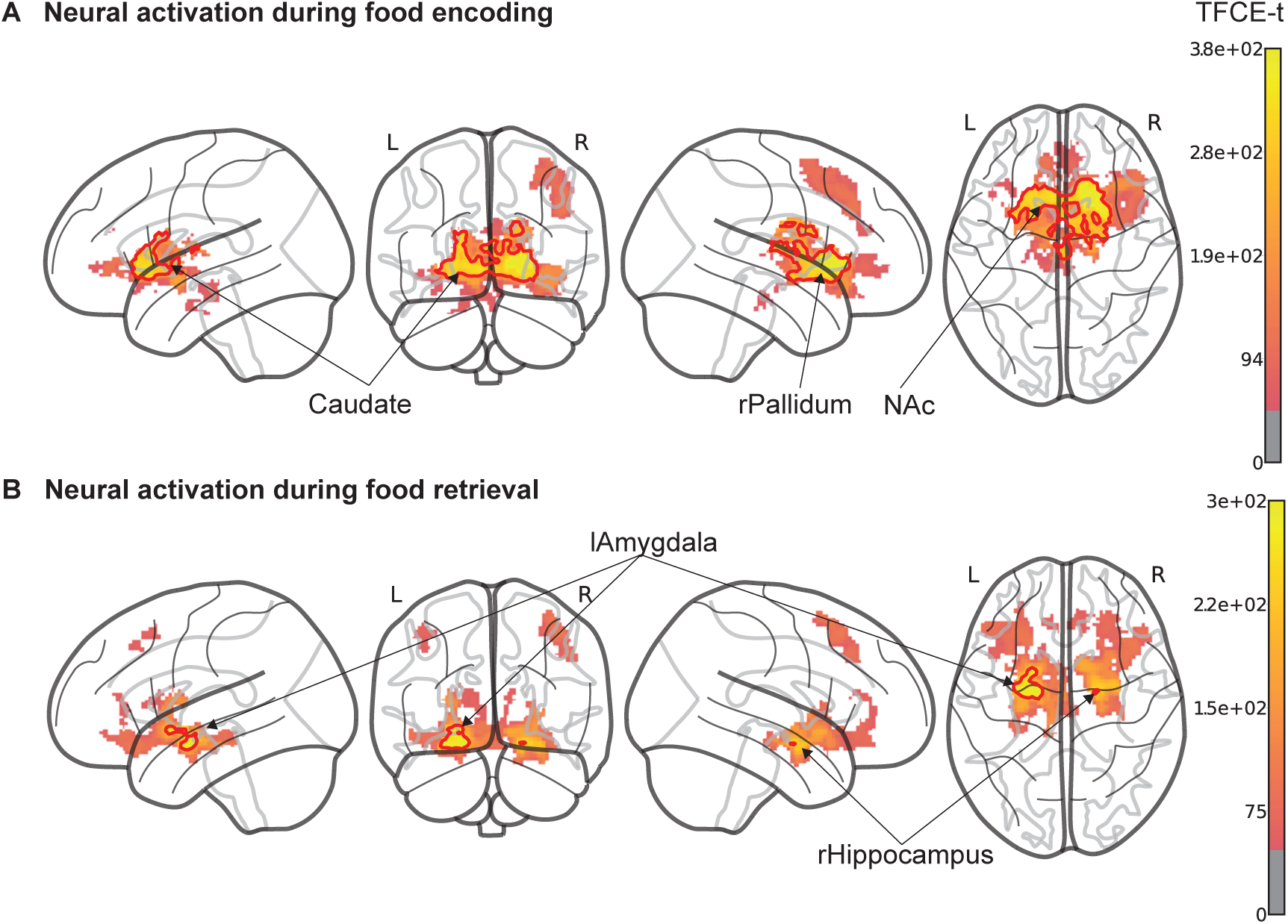
Neural response during food compared to art memory encoding and retrieval. (A) During encoding, successful recall of food compared with art stimuli elicited brain activations, particularly in the nucleus accumbens and reward areas. (B) During retrieval, food compared with art stimuli elicited brain activations in the left amygdala and right hippocampus when the 2nd level analysis was not corrected for sex. Statistics were done on voxel-wise blood-oxygen-level-dependent signal using the sandwich estimator toolbox with threshold-free cluster enhancement (TFCE) family-wise-error correction (FWE) of multiple comparisons (main analyses) and covariates of sex (for A only) and wanting of individual stimuli. Color bars depict parametric TFCE statistic (TFCE-t >50 for visualization purposes) with wild-boot strapped pFWE<0.05 marked in red outline.

Secondary analyses revealed that food retrieval elicited neural activations in the frontal gyrus, OFC, NAc, and putamen, but also in the hippocampus and amygdala (*clusters 1-2, 176-32320 mm*^3^*, total: 32496 mm*^3^*; PTFCE-FWE=0.037-0.001*). In parallel, art retrieval elicited similar activations in the frontal gyrus, OFC, and other reward-associated areas but not in the hippocampus and adjacent amygdala (*clusters 1-5, 16-19200 mm*^3^*, total: 20064 mm*^3^*; PTFCE-FWE<0.05-0.002;* Supplementary Figure 2, Supplementary Table 4). Moreover, during retrieval of old food stimuli, bilateral caudate and pallidum were activated (*clusters 1-3, 624-2568 mm*^3^*, total: 5384 mm*^3^*; PTFCE-FWE<0.027-0.004*). Considering further pre-registered analyses, direct comparison of food versus art recognition showed differences in amygdala, hippocampus, putamen, OFC, dmPFC, cingulate, hypothalamus, and brainstem activity before FWE correction (*H3_m, H2_e*; *clusters 1-26, 8-12856 mm*^3^*; total: 23160 mm*^3^*; PTFCE=0.033-0.0487; PTFCE-FWE>0.05*); and analyses without the correction for sex showed left amygdala and right hippocampus activations (*clusters 1-2, 24-800 mm*^3^*, total: 824 mm*^3^*; PTFCE-FWE=0.048-0.029*; Figure 2B, Supplementary Table 4).

### Changes in neural correlates of food memory after prebiotic intervention

To understand whether the gut-brain axis modulates food-specific memory, we next assayed participants’ interventional data from a crossover (within-subject) randomized controlled trial (RCT;^32^). Participants received either a high-dose prebiotic verum (inulin) or an equicaloric placebo (maltodextrin) daily for 14 days, followed by a 2-week wash-out period, and then received the placebo/verum (see Methods for details). Memory-fMRI was acquired at four timepoints, i.e. before and after the intervention and placebo period, respectively.

As reported using linear mixed effect modeling of within-scanner button presses^31^, the prebiotic intervention did not affect memory accuracy (Figure 3A; Supplementary Table 6). However, the prebiotic intervention compared to placebo significantly reduced neural activations in reward areas, including dmPFC, ACC, and OFC during successful food compared to art memory encoding and retrieval, respectively (Figure 3B and 3C; *H3_e*; Supplementary Table 4). Specifically, while neural activations during encoding showed centralized clusters within the anterior end of the pregenual ACC (pgACC; *clusters 1-3, 40-864 mm*^3^*, total: 1056 mm*^3^*; PTFCE-FEW<0.05-0.033*), those during retrieval extended across the ACC and bilateral dmPFC (*clusters 1-6, 336-1240 mm*^3^*, total: 4152 mm*^3^*; PTFCE-FWE=0.041-0.009*). According to post-hoc analyses, we observed during retrieval of known (old) food compared to art stimuli, decreased activations after prebiotic intervention in cortical dmPFC, ACC, and OFC, but also caudate, hippocampus, thalamus, VTA and homeostatic system associated hypothalamus compared to placebo (*clusters 1-2, 9376-24440 mm*^3^*, total: 33816 mm*^3^*; PTFCE-FEW=0.014-0.009*).

**Figure 3:**
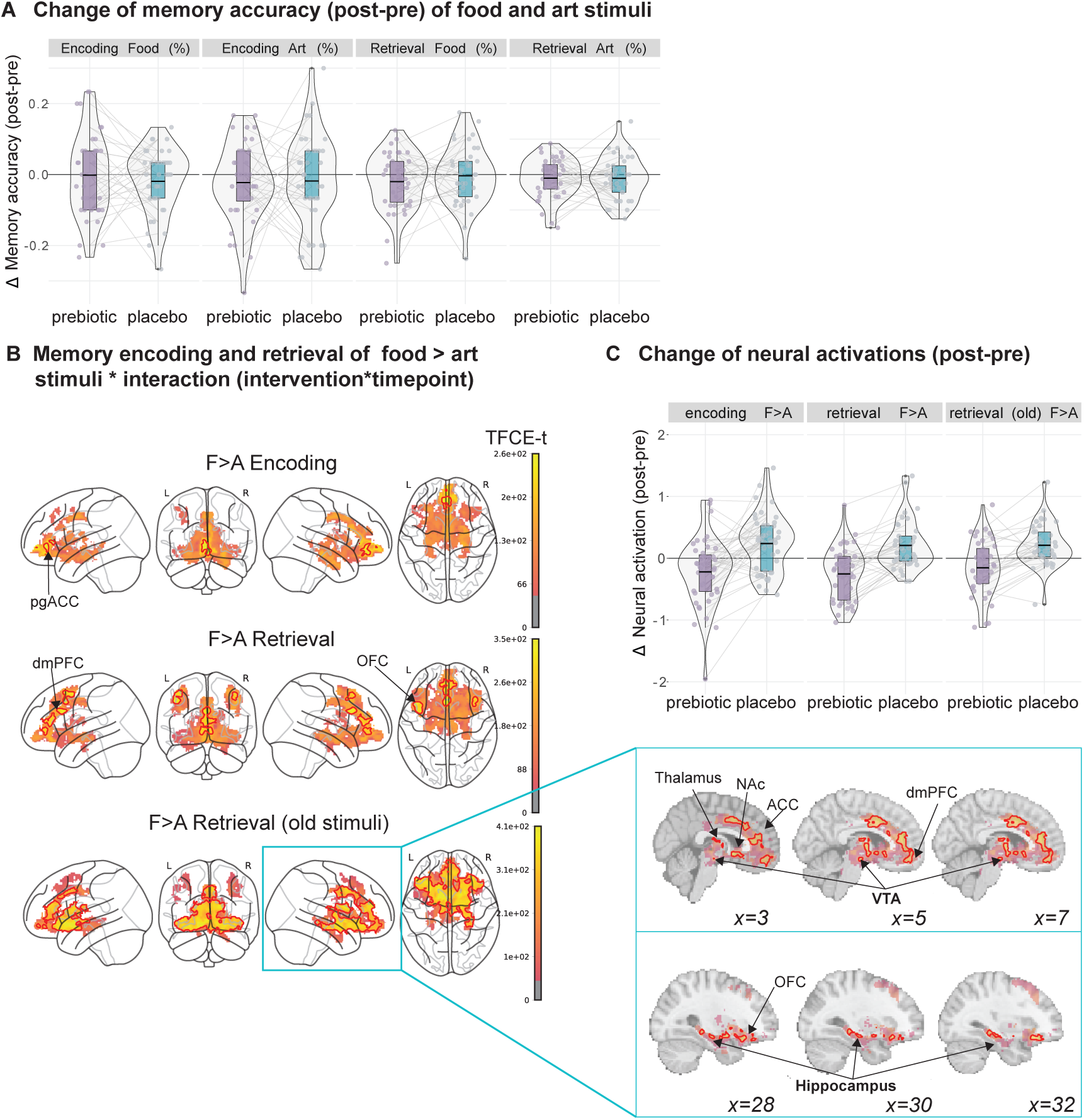
Effects of prebiotic intervention on food memory encoding and retrieval. (A) After intervention, participants’ memory performance (response accuracy) did not differ between groups (analysis of variance (ANOVA) analysis for null-full model comparisons (p<0.05), exploratory analysis, nsubj=55). (B) Food (F) compared to art (A) memory encoding (top row), retrieval (middle row) and retrieval of old stimuli (bottom row) elicited neural response in dorsomedial PFC, ACC, and OFC (F>A encoding and retrieval), and further caudate, hippocampus, thalamus, VTA, and hypothalamus (F>A retrieval of old stimuli). (C) The difference (delta) between baseline and follow-up (post-pre) for neural activations presented for prebiotic and placebo groups. The brain activation responses are shown to decrease in the prebiotic group compared to the placebo. Statistics were done according to linear mixed effects modelling, up to 4 time points per participant × 80 (encoding, retrieval old) or 160 (retrieval) stimuli on memory response accuracy and on voxel-wise blood-oxygen-level-dependent signal using the sandwich estimator toolbox with threshold-free cluster enhancement (TFCE) family-wise-error correction (FWE) of multiple comparisons and covariates of sex and wanting of individual stimuli. Color bars depict parametric TFCE statistic with wild-boot strapped pFWE<0.05 marked in red outline (upper right panel) and as an enlargement (lower right panel).

Moreover, we found that food compared to art memory encoding and retrieval correlated with reduced neural activations in the body of the hippocampus after prebiotic intervention compared to placebo (during encoding: *cluster 1, 640 mm*^3^*; PTFCE-FEW=0.015;* during retrieval: *clusters 1-2, 64-128 mm*^3^*, total: 192 mm*^3^*; PTFCE-FEW=0.0042-0.04;* Supplementary Figure 3A-C, Supplementary Table 5).

We also observed lower neural activations during encoding and retrieval after prebiotics compared to placebo when taking the stimuli’s subjective values into account (*encoding*wanting: cluster 1-14, 8-24 mm3, total: 152mm3, pTFCE=0.0135, pTFCE-FWE>0.05; retrieval*wanting: cluster 1-21, 8-1120mm3, total: 3552mm3, pTFCE=0.00302-0.0479, pTFCE-FWE>0.05*). These activations were predominantly within the amygdala, hippocampus, parahippocampus, cingulate, dmPFC, and OFC. However, those wanting-dependent differences in neural activations after intervention did not survive FWE corrections.

In sum, we show a significant prebiotic effect on the neural activations during memory encoding and retrieval independent from wanting (H3_e), which did however not reach significance when taking subjective value into account (*H4_e* & *H5_e*).

### Memory-related activity in the hippocampus and microbial and metabolic outcomes at first study visit

Next, we tested if activations of the hippocampus during successful encoding and retrieval relate to diversity and abundance of gut microbiota at the first study visit. Therefore, we identified neural activations during successful memory retrieval at the first study visit, extracted individual’s mean activations across two main clusters (Figure 4A), and run comparisons with microbial and metabolic measures derived from feces and blood using 16S rRNA sequencing and mass spectroscopy and ELISA, respectively (see Supplementary Table 8 and Methods for details). Similar to when taking all four timepoints into account, we found activations in vmPFC, subcallosal cortex, NAc, amygdala, and parahippocampus (*clusters 1-8, 8-3096 mm*^3^*, total: 6376 mm*^3^*; PTFCE-FEW=0.048-0.012;* Figure 4A; Supplementary Table 4). The neural activations of clusters number three and seven (*cluster 3*, *184 mm*^3^*; PTFCE-FEW=0.037; cluster 7*, *176 mm*^3^*; PTFCE-FEW=0.029)* during memory retrieval at the first study visit were in their peak activations within the left amygdala to parahippocampus but included some activations within the head of the hippocampus. Parameter estimates from clusters three and seven were then associated with gut microbiota and blood outcomes at first study visit. While analyses showed no associations of brain activation parameters with microbiota relative abundances after FDR correction (*H4_m*), exploratory analyses revealed that higher left parahippocampus/amygdala (cluster 3) activations correlated with higher fecal SCFA propionate (*rho=0.38, p=0.017, pFDR=0.049;* Figure 4B) and fecal SCFA butyrate (*rho=0.32, p=0.047, pFDR=0.070*), however, the latter did not survive FDR correction. Moreover, higher left parahippocampus (cluster 7) activations correlated with total SCFA levels (mg; *rho=0.38, p=0.005, pFDR=0.014*).

**Figure 4:**
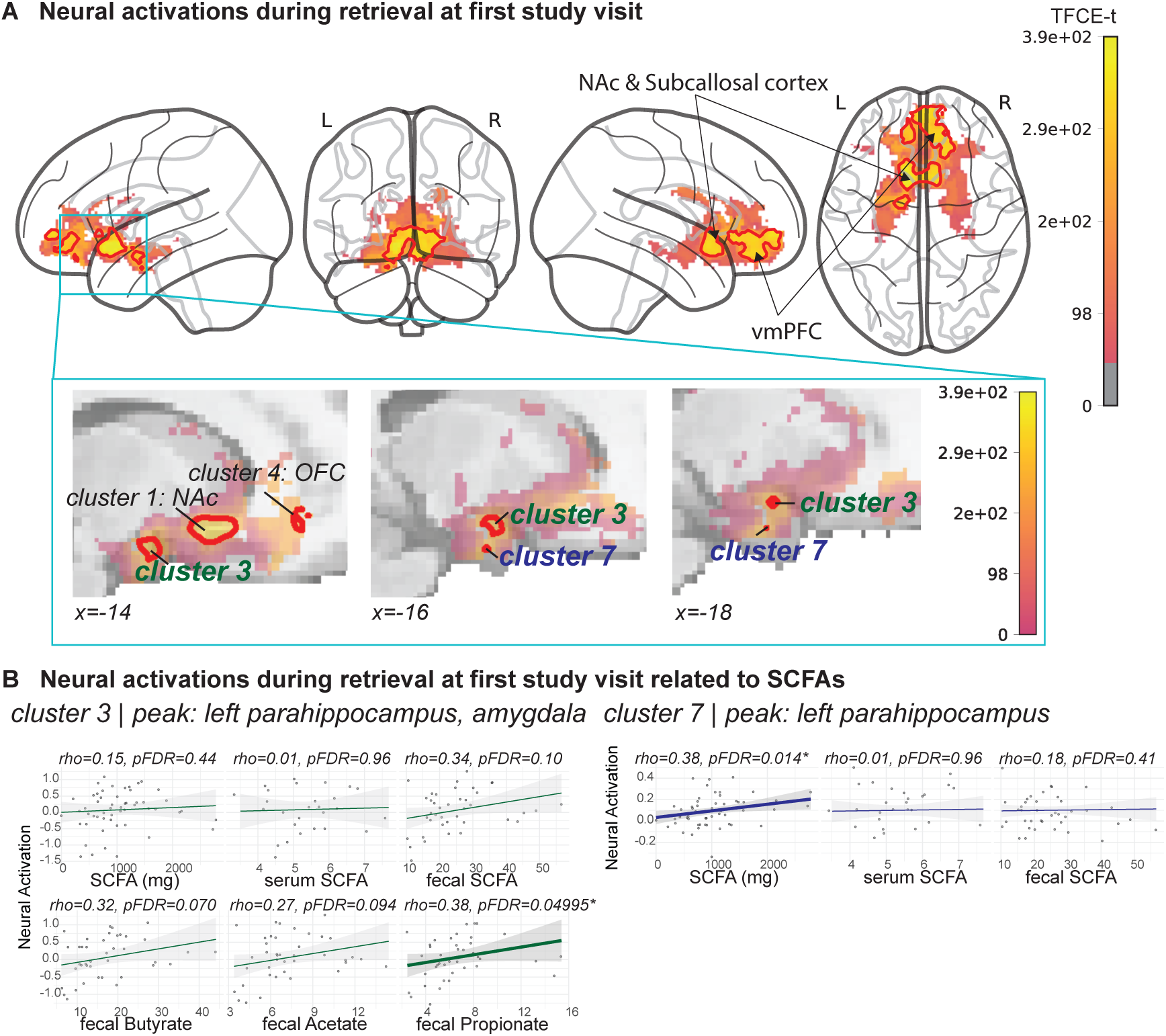
Neural activations during memory retrieval at first study visit (session 1). Memory retrieval of (A) food and art elicited neural responses in vmPFC, NAc, amygdala, parahippocampus, and hippocampus head (B). Neural activations of clusters three and seven in A are related to small-chain fatty acids (SCFAs) levels after FDR correction.

### Prebiotics-induced changes in microbial and metabolic outcomes and hippocampal activations during food-related memory

We analyzed if significant intervention-induced changes in gut microbiota (H6_e, H8_e) and metabolic outcomes (H7_e; see categories in Methods) predict intervention-induced changes in neural activations of the hippocampus during retrieval and encoding (Supplementary Table 7). Therefore, we extracted parameter estimates from neural activation of respective interaction effects (Supplementary Table 4-5).

Overall, we found that the neural response of the hippocampus during successful memory retrieval (contrast: food+art), was not related to microbial abundance measures (*H6_e*) but to a range of blood outcomes (*H7_e*; Table S9). Specifically, neural activations during retrieval, which included hippocampus and amygdala activations, were related to the liver enzyme Aspartate Aminotransferase (ASAT*; rho=-0.177, pFDR=0.041*). Further associations were revealed when using the hippocampal explicit mask (head and body of hippocampus). Here, the neural activations related to SCFA serum lactate (*rho=-0.208, pFDR=0.048*), ASAT (*rho=-0.193, pFDR=0.024*), fasting glucose levels (*rho=-0.193, pFDR=0.048*), carnitine (*rho=-0.212, pFDR=0.017*), tryptophan (TryM: *rho=-0.228, pFDR=0.017* and TryLNAA: *rho=-0.198, pFDR=0.023*). No associations emerged for vitamin levels.

However, when investigating differences in the neural correlates for food compared to art memory (contrast: food>art), hippocampal activations were related to microbial abundance measures implicated in obesity (see Supplementary Table 10). Here we found, neural activations in the hippocampus during food>art encoding (using a hippocampal mask) was related to the abundance change of *Firmicutes* (prebiotics: *rho=0.384, pFDR=0.025;* Figure 5A; *H8_e*). This was also confirmed using the hippocampus explicit mask within the equivalent contrast (prebiotic: *rho=0.421, pFDR=0.012*; Supplementary Figure 3D). Neural activations during food>art retrieval in the hippocampus (extracted parameter estimates using a hippocampal mask^40^), showed inverse association with the change in diversity *Evenness* measure after prebiotics, but not after placebo; however, this did not survive FDR correction for multiple comparisons (prebiotic: *rho=-3.376, pFDR=0.057, p=0.019;* Figure 5B).

**Figure 5:**
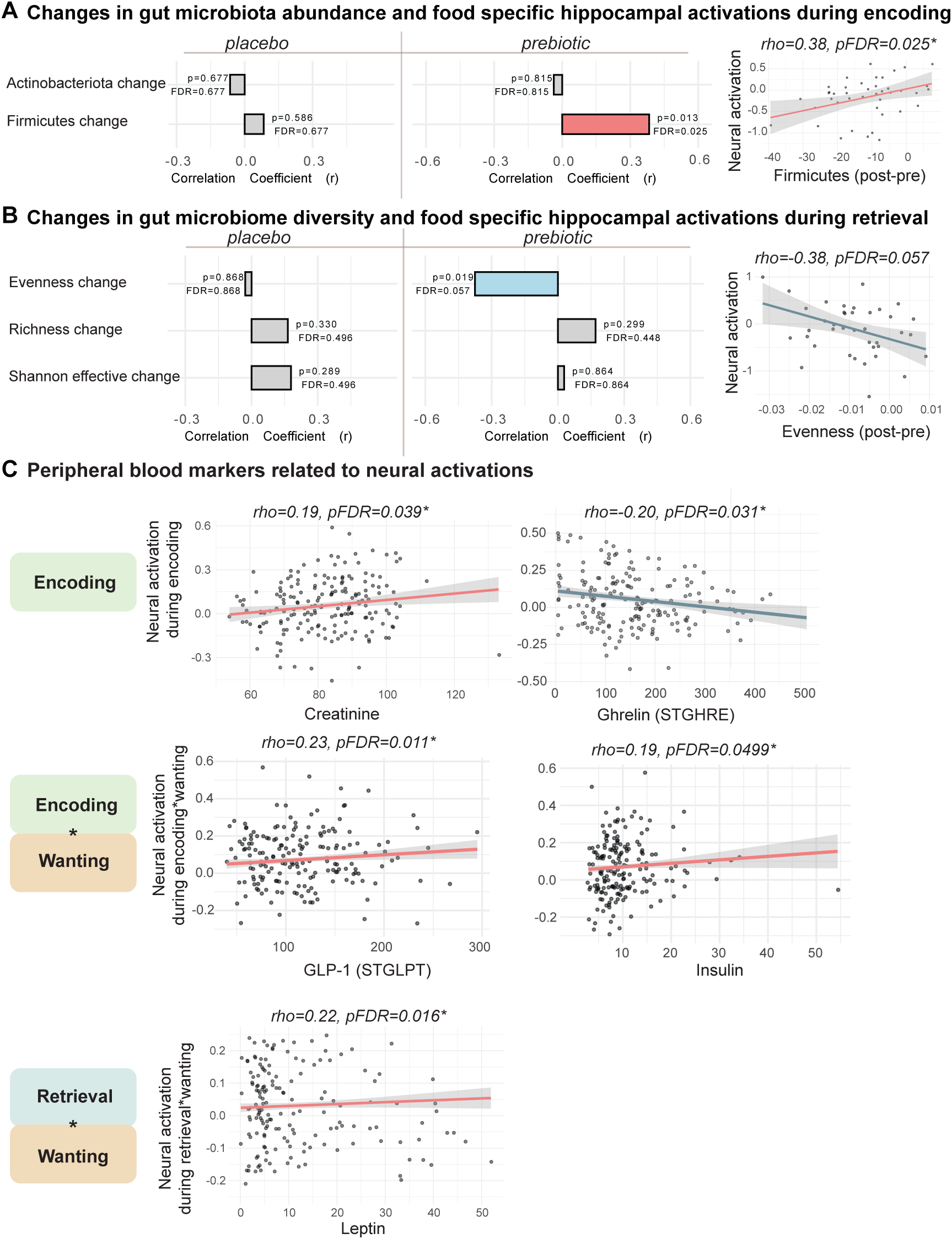
Spearman correlation between hippocampal and homeostatic-related neural response with gut microbiota, metabolic, and hormonal markers. (A) Correlation between the change of hippocampal neural activations (post-to-pre intervention difference – delta), extracted from 2nd level analysis (interaction effects), and the change of gut microbiome abundance in the phylum and (B) diversity. Data is separated by intervention groups, showing correlations in the placebo (left) and prebiotic (right) groups. Red bars indicate direct correlations, blue bars indicate inverse correlations, and grey bars are non-significant correlations using p<0.05 before FDR correction. Scatterplots on the right show sample distributions. (C) present relationships between peripheral blood markers related to neural activations (main effects) and the mean levels (across 4 sessions) during encoding (top), wanting-modulated encoding (middle), and wanting-modulated retrieval (bottom).

### Neural responses during memory encoding and retrieval of the hypothalamus relate to homeostatic outcomes

We found that neural activity in the hypothalamus during encoding (contrast: food+art) was related to creatinine levels (*rho=0.189, pFDR=0.039*), and inversely to fasting ghrelin levels (*rho=-0.202, pFDR=0.031*; Figure 5C). Neural activity during encoding modulated by wanting (see Figure 1C) was related to fasting insulin (*rho=0.172, pFDR<0.05*) and fasting GLP-1 levels (*rho=0.227, pFDR=0.011),* and neural response during retrieval modulated by wanting (see Figure 1D) was related to leptin levels (*rho=0.224, pFDR=0.016*) and TSH (*rho=0.211, pFDR=0.028*). Post-hoc Spearman correlations adjusted for BMI showed inverse relationships between ghrelin and fasting GLP-1 levels (*r=-0.31, p<0.001*), ghrelin and insulin (*r=-022, p=0.03*), GLP-1 (*r=-0.21, p=0.01)* and leptin (*r=-0.21, p<0.001)* with creatinine, and direct associations between fasting GLP-1 (*r=021, p=0.04*), and leptin (*r=0.32, p<0.001*) and insulin levels. Moreover, BMI was associated with fasting ghrelin (*r=-0.20, p<0.01*), GLP-1 (*r=0.22, p<0.01*), and insulin (*r=0.32, p<0.001)*.

### Neural responses during memory encoding and retrieval of other reward areas relate to gut microbial and blood outcomes

Exploratory analyses were conducted to test main neural responses of predominantly reward areas during encoding and retrieval (see Figure 1-2 and Supplementary Figure 1-2) with gut microbiota diversity, abundance, and blood outcome levels. All results are shown in Supplementary Table 9 and 10.

Besides the already mentioned associations between neural responses during encoding and retrieval modulated by wanting related to GLP-1, insulin, leptin, and TSH levels, respectively, we found that the neural responses of food compared to art encoding (contrast: food>art) are related to SCFA fecal acetate (*rho=-0.231, pFDR=0.011*). Additionally, during food>art encoding modulated by wanting, the neural activations (in right ACC) is related to *Actinobacteriota* abundance (*rho=0.226, pFDR=0.019*). No associations were shown with gut microbiota diversity, amino acid derivatives, vitamin levels, and liver enzyme markers. For further exploratory analyses, see Supplementary Data 12 and Supplementary Figure 4.

## Discussion

Using pre-registered analyses of task fMRI in 55 adults with overweight, we demonstrate that food memory encoding and retrieval elicited brain activations in homeostatic and hedonic areas, and that a higher subjective valuation of a given stimulus correlated with stronger brain activations in cingulate, dmPFC, VTA, and hypothalamus during encoding and retrieval. Also, food, compared to art retrieval, elicited specific activations involving the hippocampus and amygdala. Independent of the subjective wanting rating of stimuli, encoding and retrieval were associated with activations in posterior OFC, frontal pole, cingulate, caudate, right thalamus, and NAc, as well as the hypothalamus. Memory-related neural activations of the hypothalamus and/or reward centers were associated with metabolites and gut hormones, including acetate, creatinine, TSH, insulin, ghrelin, GLP-1, and leptin levels (Figure 6). Moreover, encoding of food compared to art stimuli, when modulated by subjective wanting, elicited specific activations in right ACC, which were further associated with *Actinobacteria* abundance.

**Figure 6:**
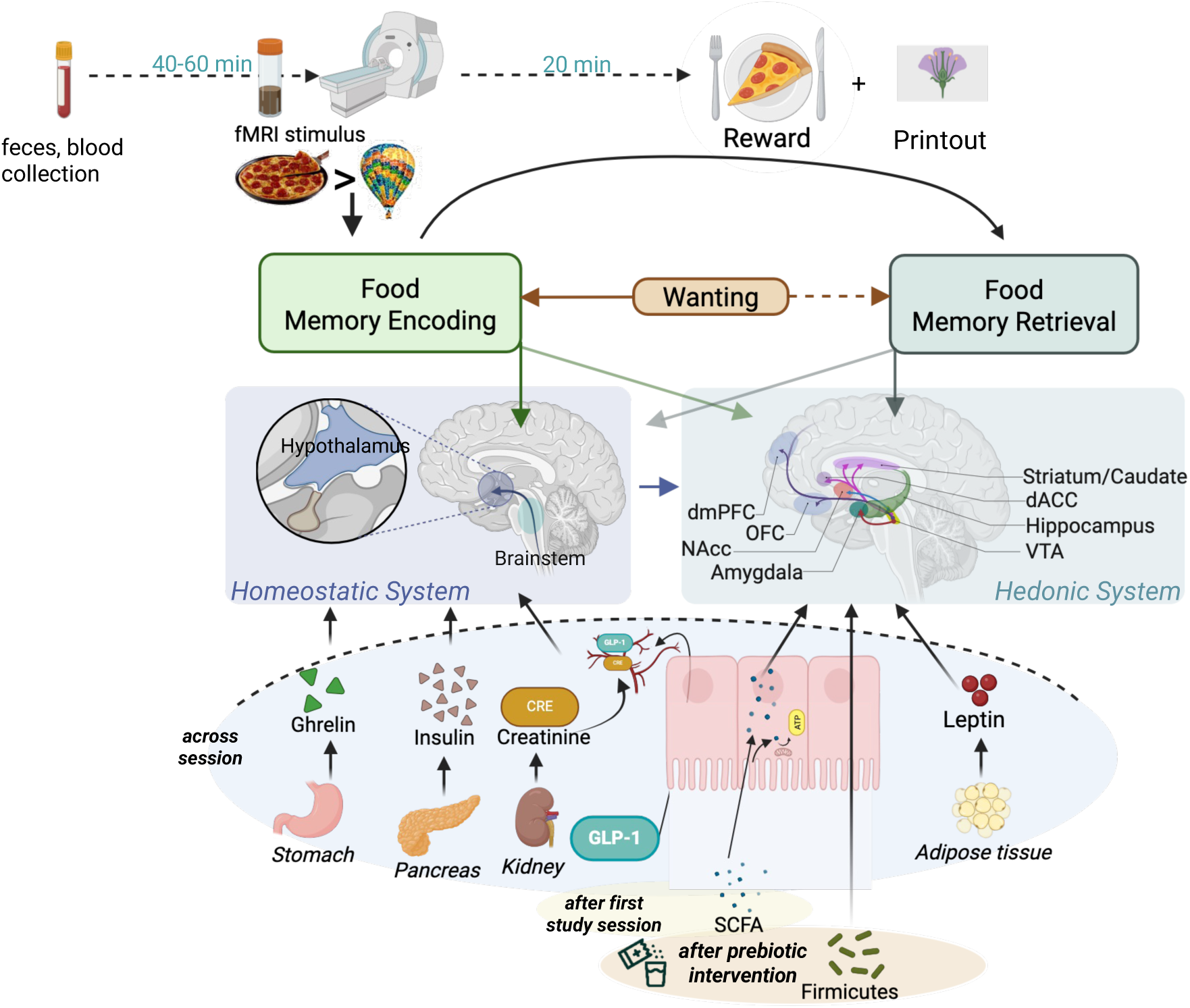
Summary of hedonic and homeostatic processes associated with neural activations during memory encoding and retrieval. Schematic overview of results presented in this study, integrating neural correlates during food versus art fMRI stimulus encoding and retrieval with activations in brain areas related to the homeostatic (hypothalamus, brainstem) and hedonic system (dmPFC, OFC, NAc, amygdala, striatum/caudate, dACC, hippocampus, and VTA). Those systems were also found to be associated with metabolites and gut hormones (ghrelin, insulin, creatine, GLP-1, and leptin) or after the first study session (SCFAs) or after prebiotic intervention (Firmicutes). Originally created in BioRender.com.

Prebiotic intervention significantly reduced neural activations during successful subsequent memory encoding and retrieval of food compared to art stimuli in homeostatic and reward regions. The reduction in hippocampal activations correlated with changes in gut microbiota composition, specifically Firmicutes abundance, and blood markers (SCFAs, glucose, carnitine, etc.). Together, these findings highlight the integration of reward, homeostatic, and metabolic signals during memory processes for food that are modifiable through dietary interventions. We show that reward-enhanced (food) memory encoding and retrieval correlated with stronger activations in reward areas, including dmPFC, VTA, and cingulum. These are likely caused by potentially higher dopaminergic signaling ^29,30^, supporting a central role of dopaminergic signaling in linking reward valuation with memory processes.

Indeed, the integration of hedonic value, such as money, has been shown previously to boost declarative memory encoding and consolidation in behavioral paradigms (reviewed in ^27^). Thus, our work aligns with and expands previous studies^27,41,42^ towards food as primary reinforcer of memory. Looking closer into wanting modulation of food compared to art stimuli, we found greater right ACC activations during encoding. This aligns with the literature as ACC is discussed as providing reward-related input into the hippocampal system via the posterior cingulate and parahippocampal gyrus^43^.

Independent of individual wanting ratings, we confirmed that neural responses during food compared to art encoding elicited more widespread activations in reward and hedonic brain regions, namely the right pallidum, caudate, and NAc. These hedonic hotspots in the brain have been extensively studied^29^, but evidence in humans on the relation to memorized food cues is limited. During subsequent memory retrieval, regardless of stimulus type, we observed activations across reward-related brain areas, including OFC, cingulate, paracingulate, and NAc. We also observed activations in “classical” memory areas such as the hippocampus and amygdala in exploratory analyses. Thus, retrieval-related activation patterns may primarily reflect memory- and emotion-related processes in the hippocampus and amygdala, supported by hedonic input. Follow-up studies are needed to confirm that the reward category (food as primary vs. art as secondary) would not distinctively predict neural correlates of memory recognition, as opposed to memory encoding according to our findings.

We also found evidence of homeostatic processes during encoding, as hypothalamic activations were observed during successful memory formation. The hypothalamus is a central structure for the regulation of food intake, integrating signals from the gut, adipose tissue, and brainstem^5,10^. This might explain the observed correlations of neural activations in the hypothalamus with peripheral hormones and metabolite concentrations (ghrelin, creatinine, TSH, insulin, GLP-1, and leptin).

Previously, ghrelin has been shown to increase hunger by stimulating the hypothalamic arcuate nucleus^10^. Also, ghrelin increased the reward from palatable food through dopaminergic signaling^44^. Surprisingly, we observed correlations between lower ghrelin levels and higher neural responses in posterior OFC, right thalamus, NAc, and hypothalamus during memory encoding. By contrast, a human fMRI study has reported increased activations after intravenous ghrelin in reward-related brain regions during food encoding^45^. Moreover, a direct enhancement of hippocampal function via hypothalamus-stimulated ghrelin modulation has been shown in mouse studies ^12,46^. Thus, future studies in humans are needed to further explore the hypothalamus-hippocampus connection related to ghrelin levels. However, we note that the estimated fasting ghrelin levels (before MRI scan) may relate differently to favorable cardiometabolic parameters and brain activations compared to studies using (acute) ghrelin infusion^46^, and estimated (postprandial) ghrelin change (i.e. area under the curve) from before to after MRI scan^47^. Thus, we suggest that higher fasting ghrelin may reflect ghrelin sensitivity improvements and further insulin sensitivity recovery, which aligns with interventional findings^48^. Moreover, this can be supported by the findings of indirect correlations between fasting ghrelin levels and insulin levels, GLP-1 levels, and BMI.

Moreover, we showed that higher neural activations during reward-related memory encoding and recognition in cingulate, dmPFC, VTA, and hypothalamus related to higher fasting insulin, GLP-1, and leptin levels. These hormones are discussed as potential biomarkers with neuroprotective effects^49^. While insulin signaling has long been established as beneficial for cognition^23,24^, the effects of GLP-1 on cognition, e.g. breakthrough effects of GLP-1 receptor agonists on eating behavior and weight loss in obesity^25^, have only recently been described.

First studies in humans indicate the potential of GLP-1 receptor agonists related to reduced risk of Alzheimer’s disease and related dementias^50^. The current results now propose that endogenous GLP-1 might help food stimuli encoding through modulation of hypothalamic input^26^. However, subcutaneous GLP-1 injection was related to reduced neural activations and wanting of high-caloric food stimuli (e.g.,^51,52^). Future studies need to confirm whether lower fasting GLP-1 levels may be beneficial for reward-related memory encoding. Moreover, we found a direct relationship between neural activity and leptin during retrieval. This finding aligns with animal literature showing impairments in hippocampal synaptic plasticity and in spatial memory performance in leptin-deficient rats^53^.

Other fasting blood markers related to metabolism and energy homeostasis, such as creatinine and TSH were shown to correlate with greater neural activations during encoding and retrieval, respectively, suggesting that optimal energy reserves may support critical neural circuits, such as the hypothalamus–hippocampus and reward networks. This is in line with a meta-analysis showing improved memory after creatine intervention^18^; however, the integrative function of TSH in hypothalamic-hippocampal pathways remains unclear^20,54,55^.

We have previously reported that dietary interventions attenuate neural responses to high-caloric, wanted food during a food preference task in the VTA and right OFC^32^. Here, we extend these findings to memory function, demonstrating reduced neural response after prebiotic intervention, compared to placebo, during food versus art retrieval in dmPFC, ACC, and OFC, and food-specific activation in the pgACC during encoding. Overall, those areas are relevant during reward-guided decision-making^56^, and in food-related memory formation. Thus, the observed reduction in neural activations in hedonic regions following prebiotic intervention suggests a modulation of reward pathways, potentially reducing the reinforcing impact of reward on food memories.

Specifically, the dmPFC encodes the structure and organization of the task environment, especially its social environment^57^, but has also been discussed to play a role in the inhibition of food-seeking responses^58^ and substance-use seeking behavior^59^. We suggest that in our study, dmPFC activations were particularly embedded in the decision-making process during memory recognition of food stimuli and were further modulated by prebiotic versus placebo supplementation.

The ACC has been implicated in food motivation and satiety^60^. The dorsal (dACC) and pregenual subregions are based on distinct functions: while dACC reflects the prospective value of a course of action required to achieve a goal ^61^, taking the time components into account ^62^; pgACC integrates costs and benefits of initiating a course of action^63^. This is in accordance with our results, as in the context of the food preference task-fMRI, we suggest that the pgACC integrated wanting of stimuli and therefore the costs and benefits of specific food stimuli during encoding. During retrieval, dACC activity potentially indicating sensitivity to the time until reward delivery (post-MRI) and timings between stimuli.

Lastly, the OFC serves as an integrative hub for orchestrating motivated food intake by integrating sensory information (i.e., taste, smell, vision; ^64^). Altered OFC function has been reported in individuals with obesity, where endophenotypes such as impaired response inhibition of food intake are linked to OFC dysfunction ^65^. Rodent studies have also shown that diets impair food intake based on the expected value ^65^. Consistent with these findings, our results demonstrate that prebiotic intervention reduced OFC activations during memory retrieval, suggesting diminished reward enhancement of food memories after prebiotic intervention.

Interestingly, when we restricted analysis to correct identification of old food versus art stimuli during retrieval, the prebiotic intervention showed a stronger association with neural responses beyond the dmPFC, ACC, and OFC. Specifically, activations in the hippocampus, caudate, thalamus, VTA, and hypothalamus were reduced after prebiotic intervention compared to placebo. Thus, accurate retrieval of previously encoded food stimuli appears to rely on a broader network of regions than typically emphasized, suggesting that memory, reward, and homeostatic circuitry jointly contribute to food-specific retrieval. These results invite further study of how these regions cooperate in food memory retrieval under metabolic modulation (Supplementary Data 13).

Moreover, our exploratory RCT analyses revealed that decreased hippocampal activation during food encoding was associated with prebiotics-induced increases in *Firmicutes* abundance, and that hippocampal changes correlated with decreases in microbiota evenness. Indeed, previous studies have reported microbiome differences between individuals with and without obesity that were related to short-term and working memory performance^66^. Consistent with our findings, this may highlight the therapeutic potential of targeting gut microbiota to mitigate memory impairment via modulation of neural activity patterns.

The following limitations should be considered. The sample was restricted to individuals with overweight, limiting generalizability to other populations. Furthermore, sex imbalance (34.5% females) and a relatively high dropout rate (∼14.5% after first study visit) can reduce statistical power and may have biased results. Another limitation is that gut hormone levels were measured in a fasted state from blood serum before fMRI.

Moreover, the design of the fMRI task resulted in a power difference between encoding and retrieval. To mitigate this imbalance, we restricted analyses at retrieval to old stimuli only and generally found stronger brain activation for the correct identification of old targets compared to including correct rejection of new stimuli. Future studies should address these limitations by including confidence ratings in the subsequent memory task.

The use of well-validated food versus art memory stimuli (food database^67,68^; art database^39^) allowed for a precise examination of neural correlates. However, memory accuracy was consistently lower for art compared to food stimuli, and motivational confounds should be considered in future designs.

Lastly, on the behavioral level, we did not observe lower memory accuracy after prebiotic intervention. Previous findings show inconsistency with an effect of four-but not eight-week prebiotic interventions on cognitive flexibility^69^ and working memory^70^, respectively. This suggests that the neural changes observed here may require longer intervention periods to manifest at the behavioral level. Yet, whether altered brain responses to food stimuli will ultimately translate into meaningful behavioral differences remains elusive based on the current data. Ongoing studies with longer intervention phases aim to clarify these effects^47^.

In conclusion, this study highlights the complex interplay between hedonic and homeostatic mechanisms in food-related memory processes and the potential of prebiotic interventions to modulate these neural processes. Our findings promote the understanding of challenges posed by modern obesogenic environments and could help to improve public health outcomes.

## Methods

This preregistered study is part of the over-arching Gut-Brain Study (https://clinicaltrials.gov/study/NCT03829189, https://osf.io/f6qz5). The design and analysis plan was preregistered at https://osf.io/whbc8.

### Participants

Participants were recruited via the institute’s database and advertisements. Inclusion criteria were a body-mass-index of 25-30 kg/m^2^, no MRI contraindications, aged 18-45 years, and women: intake of oral contraceptives. Exclusion criteria were: neurological or psychiatric disease; MRI contraindications; daily consumption of > 50g alcohol, >10 cigarettes, or >6 cups of coffee; intake of medication acting on the central nervous system; diabetes mellitus type 2 or other severe metabolic disease; severe untreated internal disease including the gastrointestinal tract, lung, heart, vasculature, liver and kidneys; eating disorder or unconventional eating habits; pregnancy, breastfeeding. After the exclusion of first test subject (see preregistration), exclusion for *a posteriori* detected neurological and psychological diseases, missed task-based functional magnetic resonance imaging (fMRI) acquisition, or too many missed trials, we included a total of 55 participants in this study.

All participants were reimbursed for participation (9-10€/h and additionally €30 for study completion) and gave written informed consent. The study was approved by the Research Ethics Committee of the University of Leipzig and was conducted in accordance with the Declaration of Helsinki. The study was registered prior to recruitment and data acquisition at https://clinicaltrials.gov/ct2/show/NCT03829189 (14/01/2019).

### Patient and public involvement

The authors acknowledge a missed opportunity of not following a tailored approach to involve patients or the public in the design of the study. We invited and collected comments and assessments from all participants throughout the study to inform the design of upcoming research studies.

### Study Design

This study is part of the Gut-Brain Study, a double-blind randomized controlled cross-over (within-subject) intervention trial study. Data acquisition took place 2019-2022. All staff members and participants were blinded regarding the study intervention/placebo allocation during data collection and data analysis, except for medical study assistants in case of monitoring of potential adverse events.

During a screening time point, state and trait characteristics of each participant were assessed. Each eligible recruited participant (N=61) was allocated to one of the two study arms, in which they received either the prebiotic verum of high-fiber inulin (30g inulin; 63 kcal, 26.7 g fiber,

Orafti Beneo Synergy1, BENEO, Mannheim, Germany) or the equicaloric placebo (16 g maltodextrin, 63 kcal, 0 g fiber) each provided as two sachets per day for 14 days, followed by an at least 2 week wash-out period, and then received placebo/verum. Before and after each intervention period, participants underwent extensive testing following identical procedures at each of the total four time points (baseline 1 and 2 – *BL1* and *BL2,* and follow-up 1 and 2 - *FU1* and *FU2*). This procedure included blood drawings, anthropometry, a standardized breakfast shake (10% of gender-individual calorie need based see Harris and Benedict, 1918), a stool sample, MRI scanning (fMRI with wanting and memory task, *T1*), cognitive testing at a computer, and questionnaires.

Participants underwent fMRI scanning as they performed a wanting and memory task using Presentation® *version 16.2 0.13.17* (Windows XP). In general, participants encoded food and art pictures during the wanting task (memory encoding) and performed free memory recall (retrieval) during the memory task.

In the wanting task, participants were shown food or art images (encoding), then asked how much they want to have the depicted food or art image and respond on an 8-point Likert scale (1 labelled as ‘not at all’ and 8 as ‘absolutely’). Participants responded using a button box by moving the cursor to the right or left of the screen. Each image stimulus was shown for 4000 ms, followed by a 4000 ms response period and a 500-4000 ms inter-stimulus interval with a 500 ms jitter until the next stimulus was presented (Figure 1A, left). Participants were previously told that they would be rewarded with the highest-rated food and art image after the MRI scan. The reward was given as a dish to eat right away and as a carton-based art print to take home.

In the memory task, participants were shown food and art images; however, some images were already shown during the wanting task. They were asked to respond using a button box with buttons pertaining to the response position on the screen, if they had seen the presented food or art image in the previous task (“old”) or if they had not seen it before (“new”; Figure 1A, right. Novel images included also similar images (lure images). Each image was shown for 500 ms and followed by a fixation cross for 0-3500 ms, a response time up to 4000 ms, and response feedback for 500 ms, and an inter-stimulus interval of 500-4000 ms.

In the wanting and memory task, participants were presented with four sets of food and art images across four sessions in randomized order. There was no repetition of image across sessions within subjects. In each task, 160 stimuli (80 food with 20 per calorie quartile and 80 art stimuli with 10 per art style) were shown, divided into 3 blocks with two breaks of 30 seconds per block. In the wanting task, the 80 art stimuli consist of 10 per stimulus type, namely 1 plant, 2 animal, and 7 object stimuli); no more than 3 images per category in a row. In the memory task, 80 art images consisted of 12 plant, 12 animal, and 56 object images with 30 old stimuli (targets), 30 similar (lures), 20 new (novels) stimuli per category (food, art). Food pictures were taken from the food-pics database^67,68^, and the art.pics database^39^ served as the source for the art stimuli.

### MRI data acquisition and preprocessing

MRI was conducted at a 3 Tesla Prisma Fit Magnetom (Siemens, Erlangen, Germany) with a 32-channel head coil. Task fMRI in an event-related design assessing memory encoding and retrieval for food and art, respectively, was acquired using T2*-weighted images: fast echo-planar-imaging (EPI) blood oxygen level-dependent (BOLD): repetition time TR = 2000ms, echo time TE = 23.60ms, flip angle FA = 80°C, field-of-view FOV = (204 mm)^2^; voxel size 2 x 2 x 2 mm^3^; gap 0.26mm; 60 slices; orientation T>C -15°; multi-band = 3, interleaved. Field maps and ap/pa were acquired to be used for correcting scanner inhomogeneities in the preprocessing pipeline.

All fMRI preprocessing was done using fMRIPREP (*version 1.2.5*^71^) and followed by quality control using visual examination and MRIQC^72^ to check for severe brain pathology (based on T1w/FLAIR image, done by study doctors) and artefacts, correct brainmask extraction, correct surface reconstruction, correct co-registration between fieldmap and EPI and EPI to T1 images. FMRI images of low quality (i.e., erroneous co-registration of EPI to T1w) were excluded from analyses.

Smoothing with FWHM kernel of 6mm was done in Statistical Parametric Mapping (SPM) *V.12.7486*. Slice Timing was not used. Framewise displacement (FD in mm) and excessive motion outliers (TR>0.9mm) were extracted with *fsl_motion_outliers*; DVARS; three region-wise global signals (CSF, GM, WM); 6 rigid-body motion parameters (3 translations and 3 rotation); TRs with excessive motion (FD>0.9mm according to Siegel et al., 2014) will be regressed out from the fMRI analysis only (by including an additional regressor labelling those TRs with 1, using fsl_motion_outliers). Timepoints in which participants showed no interest in the stimuli at all (if all stimuli are ranked 1 out 8 for either food or art; was the case for 1 person at 2 timepoints) will be excluded from the SPM analysis as the parametric modulator is collinear to the onset regressor.

Moreover, we checked for signal drop-out and temporal signal-to-noise ratio (tSNR) of brain masks in region-of-interest (ROI) areas (see defined masks below).

Missed trials during the wanting and memory task fMRI (i.e., missed response): will be handled in two ways according to the randomness of trials. If missed trials occur in blocks of at least 3 stimuli missed in a row, those are considered non-random and likely due to inattentiveness or sleep of the participants (occurred in 0 (encoding) and 20 (retrieval) participant-session cases). 10% of all 160 stimuli were missed in 0 (encoding) and 17 (retrieval) participant-session cases. Those missed trials were modelled as separate regressors in fMRI analyses. In contrast, missed trials scattered in time may be randomly missed and thus are imputed using the individual average of the concurrent memory retrieval button press or wanting rating (for stimuli shown in the memory task only). This was done to allow inferences about BOLD activity during picture evaluation and has already been corrected for the wanting task^32^. For the memory task this occurred in 0 (retrieval) participant-session cases. Imputation of single-memory decision was done if missed stimuli were up to 10% of all stimuli in one session for memory retrieval, respectively, and replaced with the individual average (occurred/imputation in 3 (retrieval) participant-session cases). (see participant information in pre-registration https://osf.io/whbc8).

### Regions of Interest

*A priori* defined ROI masks were created using *fslmaths* in FMRIB Software Library (FSL; *version 6.0.3***73**) and bilateral binary masks were then used in voxel-wise fMRI analysis. The primary mask included regions related to homeostatic and hedonic regulation of food intake, eating behavior, and cognitive reward centers. As the hypothalamus plays a major role in the control of appetite by processing afferent signals from the gut and brainstem^10^, we merged the previously used primary mask^32^ based on a neurosynch.org meta-analysis of studies including keywords “hypothalamus” and “reward” (thresholded at zmin=1.96; alpha<0.05, uncorr.) with ROIs of the hypothalamus^74^ (threshold of z=10), NAc, locus coeruleus, hypothalamus, and amygdala (previously used in ^74,75^), dorsolateral prefrontal cortex, orbitofrontal cortex, anterior cingulate cortex, entorhinal cortex (Harvard-Oxford cortical atlas), hippocampus head, body and tail masks^40^, and ventral tegmental area (AAN atlas, subsampled to 2mm and threshold with 0.2).

### Caloric bias

The nutrient values, including the calorie content, were taken from the food-pics database^67,68^. The respective fiber content was assessed within control analyses. We further calculated the memory response accuracy during encoding and retrieval for food images, which are categorized in caloric quantiles (cal 1-4) and ten food types (see ^32^).

### Blood and feces markers

Fecal markers were assessed, including stool samples taken within 1–2 days before the testing day, and gut microbiota were analyzed using 16S rRNA analysis (see ^32^). Blood draw was obtained in fasting state (12.5±2.2 hours fasted) at the same time per participant for each session. Feces and blood markers were obtained to assess markers within the following categories:

- gut microbial markers: gut microbiota diversity measures: Stool frequency, Evenness, Richness, Shannon effective (i.e. measure of alpha diversity); abundance of microbiota using the abundance in their phylum (i.e. *Actinobacteriota, Bacteroidota, Fusobacteriota, Firmicutes, Proteobacteria, Verrumcumicrobiota, Cyanobacteria, Patescibacteria*), the ratio of *Bacteroidota* to *Firmicutes* (B/F ratio) and abundance in gut microbiota genera (e.g. *Bifidobacterium*) and family (e.g., *Bifidobacteriaceae*)
- fecal and serum small-chain fatty acids (SCFA) levels (i.e. total, Butyrate, Acetate, Propionate, Lactate)
- Inflammation (high-reactive C-reactive protein (hsCRP), tumor necrosis factor α (TNF-α), trimethylamine N-oxide (TMAO), TSH, interleukin-6 (IL6)),
- metabolic indicators (Glucose, Creatinine, GRF_CKDEPI_Creatinine, GFR.MDRD),
- metabolic pathways (Pyruvate, Cysteine and methionine, Flavonoid biosynthesis, Purine, Methane, Arginine and proline, Selene compound)
- gut hormones (Leptin, Insulin, Ghrelin, GLP-1, PYY),
- amino acids derivatives (Betain, Cholin, Carnitin, TyrM, TyrLNAA, TrpM),
- liver enzymes (Aspartate Aminotransferase (ASAT), Alanine Aminotransferase (ALAT)).

### Covariates

Behavioral assessments included as covariates in the statistical models were acquired at each timepoint. These include demographic data (age, socioeconomic status index measured using the mean measures of degree, occupation, and income), gender according to a questionnaire (options: male, female, diverse, no answer). Following the pre-registration, we included in all fMRI Model contrasts, gender (male, female), six rigid motion parameters, and binary regressor for TRs exceeding a motion threshold of 0.9 mm. As first results showed a high influence of the wanting category on memory response accuracy, we decided to include individual wanting ratings as an additional confounder. We included the rated ‘complexity’ of the task as well as individual stimuli as confounds during sensitivity analysis.

### Statistical Analysis

#### Behavioral tests were conducted in *R*

Memory accuracy for food and art images in four sessions was measured using the total number of hits of targets (i.e., old images) for encoding and retrieval, and correct rejection of novel or lures (i.e., new or similar images) for retrieval, divided by the possible correct memory decision. Thus, 60 targets were presented during encoding in the wanting task, and 160 targets and novels were presented during memory retrieval during the memory task.

All included 55 subjects performed above chance at food and art memory encoding as well as food memory retrieval, but only 17 subjects performed better than chance; thus, 33 performed lower with 0.46+-0.02% correct art retrieval judgements (Figure 1B, differences in their mean were estimated using t-tests). Descriptive statistics for memory accuracy are reported in Table S2.

For the MRI analysis, all voxel-wise fMRI analyses were conducted with SPM, run under *MATLAB R2021a, version 9.10*. For further ROI and microbiome analyses and image creation, we used *Python version 2.7.18*, *R version 6.11* (in *RStudio version 2024.04.2+764*), and FSL *version 6.0.3*.

The fMRI data were analyzed at two levels: At the individual level, a separate GLM was constructed for each participant. Here, the onsets of food and non-food stimuli presentation were modelled as separate regressors convolving delta functions with a canonical hemodynamic response function. In addition, combined rating scores were entered as parametric modulators of food and non-food regressors separately. Parameter estimates from events of interest were then carried forward to the second-level random effects factorial ANOVAs to test for group-level effects.

Encoding memory is tracked using the EPIs from the wanting-task 0-4000ms for those pictures which were shown during the memory task. We captured memory retrieval by using the first 4000ms of the memory task. Retrieval is thought to happen while they first see the picture (old, new, similar), before they decide on ‘new’ or ‘old’ and before button press. Note, the duration of 4000ms for pictures is modelled in SPM directly (duration of two TRs from picture onset to onset of rating phase). 6 motion regressors (not binary TR regressor in this example, as no TR surpassed 0.9 mm FD) are modelled but not tested in all models.

As preregistered, first-level contrasts of interest estimated the global difference between food and art viewing during successful encoding of stimuli in the wanting task (stimuli which were shown in the memory task) and during memory retrieval in the memory task (designs: encoding/retrieval), and the modulation of the memory performance by wanting of the stimuli (designs: encoding_wanting/retrieval_wanting), or the caloric density for food memory (designs: encoding/retrieval_kcal and encoding_wantingkcal; details on model designs in Supplementary Data 14).

Inference tests were performed on first-level contrasts and second-level factors time (baseline, follow-up), group (prebiotics/placebo), and time*group interactions, using the Sandwich Estimator (SwE) V.2.2.2^76^, implemented in SPM run in MATLAB and R. In all analyses, implicit masks with a threshold of 0.4 during 1^st^-level analysis and explicit mask ROI brain mask (defined in ROI method section) were applied during 2^nd^-level analyses. Within post-hoc analyses, main analyses were repeated using an explicit mask of the hippocampus mask (including head, body and tail sub-region^40^).

Significant results were reported according to threshold-free cluster enhancement (TFCE) methods with alpha<0.05, and non-parametric bootstrapping with family-wise error (FWE) correction for multiple comparisons (TFCE-p-FWE<0.05). Parameter estimates of effects (TFCE-p-FWE<0.05) were extracted from global effects and specific clusters including ROIs for specific hypotheses (memory-related hippocampus and homeostatic hypothalamus).

### Neural activations associated with blood and feces markers

For addressing hypotheses about changes in neurobehavioral measures related or mediated by gut microbiome and assessed blood markers (H4_m, H6_e, H7_e, H8_e), we used aggregated neural data (using extracted individual parameter estimates of neural activations from 2^nd^-level analyses, see above) and relate those to changes to gut microbiota and blood draw outcomes using bivariate Spearman correlation analyses (using the corr() function for correlation coefficient).

First, we assessed the main neural activation parameter estimates of the hippocampus at the first study visit (ses-01) and across all four sessions related to all categories of microbiota and blood markers (H4_m).

Next, correlation analyses were done on the difference (delta) post versus pre after intervention, in those microbiota and blood outcomes that showed a significant group*time interaction effect using mixed effects inference with (restricted) maximum likelihood fitting and χ2 test for comparison of a model-of-interest and a null model for each effect of interest,

Model residuals were tested for normal distribution using the R package *performance()* with the command *check_normality(x,effects=’random’)*].

Post-hoc analyses also included analyses using neural activations (main, and interaction) shown within the hypothalamus associated with mean (across 4 sessions and prebiotic/placebo groups) and delta (post-pre after intervention) of homeostatic outcomes.

For exploratory mediation analyses, statistical inference was tested using linear models using *lmer()* in R for significant interactions identified in previous Spearman correlation analyses. Only if all paths showed significance, we tested potential mediation effects.

Results are considered significant after applying a threshold of *alpha<0.05* and false discovery rate (FDR) correction for multiple comparisons within each measure category (see methods ‘blood and feces markers’).

### Control analyses

To account for differences in false response rates between food and art, we conducted control analyses to monitor potential response biases between correct and incorrect responses, with higher values reflecting a stronger bias towards correct responses.

To ensure adequate signal quality, we calculated the tSNR for ROI across all sessions in the memory task, with equivalent measurements conducted during the wanting task (see Supplementary Figure 1).

## Data availability

The original data are accessible on OSF https://osf.io/whbc8/files. Statistical MRI maps are available at Neurovault https://neurovault.org/collections/22037/.

## Code availability

The code used in this study has been made publicly available at the following GitHub repository: https://github.com/dariajensen/memory-tfmri.

## Acknowledgments

We are very thankful to Anne-Katrin Brecht, Larissa de Biasi, Leonie Disch, Lina Eisenberg, Silke Friedrich, Laura Hesse, Ramona Menger, Maria Paerisch, Niklas Hlubek, Lynn Mosesku, Lukas Recker, Lennard Schneidewind, Emira Shehabi, Hannah Stock, Christian Schneider, Emmy Töws, Anna-Luisa Wehle, Charlotte Wiegank and Marie Zedler who were involved in the recruitment of participants, data collection as well as helping with the organising and preprocessing of the data. **DEAJ** was supported by the German Research Foundation (Deutsche Forschungsgemeinschaft, DFG; project number 209933838, CRC 1052/3, subproject A01 and CRC 931900-050), Saxon State Ministry of Science and Cultural Affairs, Projekt: LIFE017-Long Covid (SAB, 100590200), and ERA4Health - Nutribrain (Projekt: “MiHealthyDiet”). **EM** was supported by Berlin School of Mind and Brain and the German Foundation for Environment. **MR** was supported by DFG – project number 209933838, CRC1052/A9. **MvB** was supported by CRC1382, project 05 (Gut-Liver-Axis) and Novo Nordisk Foundation (Grant NNF21OC0066551). URK and MvB declare that funding was received from the ProMetheus platform for proteomics and metabolomics as part of the major infrastructure initiative CITEPro (Chemicals in the Terrestrial Environment Profiler) funded by the Helmholtz Association. **FB** was supported by the DFG (Walther-Benjamin program, project number: 464596826). The inulin supplement was sponsored by the manufacturer BENEO, Mannheim, Germany. **VW** was supported by the DFG (project number 209933838, CRC 1052/3, subproject A01 and CRC 931900-050), and ERA4Health - Nutribrain (Projekt: “MiHealthyDiet”).

## Contributions

Daria E. A. Jensen: Conceptualization, Methodology, Software, Formal analysis, Writing - Original Draft, Writing - Review & Editing, Data Curation, Visualization

Ronja Thieleking: Conceptualization, Methodology

Evelyn Medawar: Methodology, Resources, Writing - Review & Editing

Madlen Reinicke: Formal analysis (SCFAs)

Ulrike Rolle-Kampczyk: Formal analysis (Microbiome data)

Martin von Bergen: Formal analysis, Funding acquisition (Microbiome data)

Michael Stumvoll: Funding acquisition

Arno Villringer: Funding acquisition

Frauke Beyer: Methodology, Writing - Review & Editing

Veronica Witte: Conceptualization, Methodology, Writing - Review & Editing, Supervision, Validation, Funding acquisition

## References

1. Higgs, S., and Spetter, M.S. (2018). Cognitive Control of Eating: the Role of Memory in Appetite and Weight Gain. Current obesity reports 7, 50–59. 10.1007/s13679-018-0296-9.

2. Higgs, S. (2005). Memory and its role in appetite regulation. Physiology and Behavior 85, 67–72. 10.1016/j.physbeh.2005.04.003.

3. Kumar, S., Higgs, S., Rutters, F., and Humphreys, G.W. (2016). Biased towards food: Electrophysiological evidence for biased attention to food stimuli. Brain and Cognition 110, 85–93. 10.1016/j.bandc.2016.04.007.

4. de Vries, R., Boesveldt, S., and de Vet, E. (2021). Locating calories: Does the high-calorie bias in human spatial memory influence how we navigate the modern food environment? Food Quality and Preference 94, 104338. 10.1016/j.foodqual.2021.104338.

5. Tulloch, A.J., Murray, S., Vaicekonyte, R., and Avena, N.M. (2015). Neural Responses to Macronutrients: Hedonic and Homeostatic Mechanisms. Gastroenterology 148, 1205–1218. 10.1053/j.gastro.2014.12.058.

6. Markowitsch, H., Risius, U., Staniloiu, A., Piefke, M., Maderwald, S., Schulte, F., and Brand, M. (2013). Retrieval, Monitoring, and Control Processes: A 7 Tesla fMRI Approach to Memory Accuracy. Frontiers in Behavioral Neuroscience 7.

7. Stark, C.E.L., and Squire, L.R. (2000). Functional Magnetic Resonance Imaging (fMRI) Activity in the Hippocampal Region during Recognition Memory. J. Neurosci. 20, 7776–7781. 10.1523/JNEUROSCI.20-20-07776.2000.

8. Kanoski, S.E., and Grill, H.J. (2017). Hippocampus Contributions to Food Intake Control: Mnemonic, Neuroanatomical, and Endocrine Mechanisms. Biol Psychiatry 81, 748–756. 10.1016/j.biopsych.2015.09.011.

9. Leng, X., Huang, Y., Zhao, S., Jiang, X., Shi, P., and Chen, H. (2022). Altered neural correlates of episodic memory for food and non-food cues in females with overweight/obesity. Appetite 175, 106074. 10.1016/j.appet.2022.106074.

10. Nicoletti, C.F., Delfino, H.B.P., Ferreira, F.C., Pinhel, M.A. de S., and Nonino, C.B. (2019). Role of eating disorders-related polymorphisms in obesity pathophysiology. Rev Endocr Metab Disord 20, 115–125. 10.1007/s11154-019-09489-w.

11. Tschöp, M., Smiley, D.L., and Heiman, M.L. (2000). Ghrelin induces adiposity in rodents. Nature 407, 908–913. 10.1038/35038090.

12. Diano, S., Farr, S.A., Benoit, S.C., McNay, E.C., da Silva, I., Horvath, B., Gaskin, F.S., Nonaka, N., Jaeger, L.B., Banks, W.A., et al. (2006). Ghrelin controls hippocampal spine synapse density and memory performance. Nat Neurosci 9, 381–388. 10.1038/nn1656.

13. Batterham, R.L., Cowley, M.A., Small, C.J., Herzog, H., Cohen, M.A., Dakin, C.L., Wren, A.M., Brynes, A.E., Low, M.J., Ghatei, M.A., et al. (2002). Gut hormone PYY3-36 physiologically inhibits food intake. Nature 418, 650–654. 10.1038/nature00887.

14. Turton, M.D., O’Shea, D., Gunn, I., Beak, S.A., Edwards, C.M.B., Meeran, K., Choi, S.J., Taylor, G.M., Heath, M.M., Lambert, P.D., et al. (1996). A role for glucagon-like peptide-1 in the central regulation of feeding. Nature 379, 69–72. 10.1038/379069a0.

15. Woods, S.C., Lotter, E.C., McKay, L.D., and Porte, D. (1979). Chronic intracerebroventricular infusion of insulin reduces food intake and body weight of baboons. Nature 282, 503–505. 10.1038/282503a0.

16. Grill, H.J. (2010). Leptin and the systems neuroscience of meal size control. Frontiers in Neuroendocrinology 31, 61–78. 10.1016/j.yfrne.2009.10.005.

17. Harvey, J. (2007). Leptin regulation of neuronal excitability and cognitive function. Curr Opin Pharmacol 7, 643–647. 10.1016/j.coph.2007.10.006.

18. Prokopidis, K., Giannos, P., Triantafyllidis, K.K., Kechagias, K.S., Forbes, S.C., and Candow, D.G. (2023). Effects of creatine supplementation on memory in healthy individuals: a systematic review and meta-analysis of randomized controlled trials. Nutrition Reviews 81, 416–427. 10.1093/nutrit/nuac064.

19. Joseph-Bravo, P., Jaimes-Hoy, L., and Charli, J.-L. (2016). Advances in TRH signaling. Rev Endocr Metab Disord 17, 545–558. 10.1007/s11154-016-9375-y.

20. Daghighi, M.-H., Poureisa, M., Ahmadi, P., Reshadatjoo, M., Golestani, S., Naghavi-Behzad, M., and Karkon-Shayan, F. (2016). Serum thyroid-stimulating hormone level and relation with size of hippocampus in patients with mild cognitive disorders. Niger Med J 57, 353–356. 10.4103/0300-1652.193862.

21. Wang, Y., Liu, Y., Yang, H., Mo, Z., Liu, H., and Yu, Q. (2025). Genetic evidence suggests a causal relationship linking thyroid function to prospective memory and dementia and Parkinson’s disease. Sci Rep 15, 20425. 10.1038/s41598-025-01596-w.

22. Li, W., Yue, L., Sun, L., and Xiao, S. (2022). An Increased Aspartate to Alanine Aminotransferase Ratio Is Associated With a Higher Risk of Cognitive Impairment. Front. Med. 9. 10.3389/fmed.2022.780174.

23. Alberry, B., and Silveira, P.P. (2023). Brain insulin signaling as a potential mediator of early life adversity effects on physical and mental health. Neuroscience & Biobehavioral Reviews 153, 105350. 10.1016/j.neubiorev.2023.105350.

24. Banks, W.A., Owen, J.B., and Erickson, M.A. (2012). Insulin in the brain: there and back again. Pharmacol Ther 136, 82–93. 10.1016/j.pharmthera.2012.07.006.

25. De Giorgi, R., Ghenciulescu, A., Dziwisz, O., Taquet, M., Adler, A.I., Koychev, I., Upthegrove, R., Solmi, M., McCutcheon, R., Pillinger, T., et al. (2025). An analysis on the role of glucagon-like peptide-1 receptor agonists in cognitive and mental health disorders. Nat. Mental Health 3, 354–373. 10.1038/s44220-025-00390-x.

26. Badulescu, S., Tabassum, A., Le, G.H., Wong, S., Phan, L., Gill, H., Llach, C.-D., McIntyre, R.S., Rosenblat, J., and Mansur, R. (2024). Glucagon-like peptide 1 agonist and effects on reward behaviour: A systematic review. Physiol Behav 283, 114622. 10.1016/j.physbeh.2024.114622.

27. Miendlarzewska, E.A., Bavelier, D., and Schwartz, S. (2016). Influence of reward motivation on human declarative memory. Neurosci Biobehav Rev 61, 156–176. 10.1016/j.neubiorev.2015.11.015.

28. Coray, R., and Quednow, B.B. (2022). The role of serotonin in declarative memory: A systematic review of animal and human research. Neuroscience & Biobehavioral Reviews 139, 104729. 10.1016/j.neubiorev.2022.104729.

29. Berridge, K.C., and Kringelbach, M.L. (2008). Affective neuroscience of pleasure: Reward in humans and animals. Psychopharmacology 199, 457–480. 10.1007/s00213-008-1099-6.

30. Kelley, A., Baldo, B., Pratt, W., and Will, M. (2005). Corticostriatal-hypothalamic circuitry and food motivation: Integration of energy, action and reward. Physiology & Behavior 86, 773–795. 10.1016/j.physbeh.2005.08.066.

31. Thieleking, R., Medawar, E., Villringer, A., Beyer, F., and Witte, A.V. (2023). Neurocognitive predictors of food memory in healthy adults – A preregistered analysis. Neurobiology of Learning and Memory 205, 107813. 10.1016/j.nlm.2023.107813.

32. Medawar, E., Beyer, F., Thieleking, R., Haange, S.-B., Rolle-Kampczyk, U., Reinicke, M., Chakaroun, R., Bergen, M. von, Stumvoll, M., Villringer, A., et al. (2024). Prebiotic diet changes neural correlates of food decision-making in overweight adults: a randomised controlled within-subject cross-over trial. Gut 73, 298–310. 10.1136/gutjnl-2023-330365.

33. Cryan, J.F., O’Riordan, K.J., Cowan, C.S.M., Sandhu, K.V., Bastiaanssen, T.F.S., Boehme, M., Codagnone, M.G., Cussotto, S., Fulling, C., Golubeva, A.V., et al. (2019). The Microbiota-Gut-Brain Axis. Physiological Reviews 99, 1877–2013. 10.1152/physrev.00018.2018.

34. Silva, Y.P., Bernardi, A., and Frozza, R.L. (2020). The Role of Short-Chain Fatty Acids From Gut Microbiota in Gut-Brain Communication. Front. Endocrinol. 11. 10.3389/fendo.2020.00025.

35. Cani, P.D., Dewever, C., and Delzenne, N.M. (2004). Inulin-type fructans modulate gastrointestinal peptides involved in appetite regulation (glucagon-like peptide-1 and ghrelin) in rats. Br J Nutr 92, 521–526. 10.1079/bjn20041225.

36. Hiel, S., Gianfrancesco, M.A., Rodriguez, J., Portheault, D., Leyrolle, Q., Bindels, L.B., Gomes Da Silveira Cauduro, C., Mulders, M.D.G.H., Zamariola, G., Azzi, A.-S., et al. (2020). Link between gut microbiota and health outcomes in inulin -treated obese patients: Lessons from the Food4Gut multicenter randomized placebo-controlled trial. Clinical Nutrition 39, 3618–3628. 10.1016/j.clnu.2020.04.005.

37. Pearce, A.L., Cevallos, M.C., Romano, O., Daoud, E., and Keller, K.L. (2022). Child meal microstructure and eating behaviors: A systematic review. Appetite 168, 105752. 10.1016/j.appet.2021.105752.

38. Stark, S.M., Kirwan, C.B., and Stark, C.E.L. (2019). Mnemonic Similarity Task: A Tool for Assessing Hippocampal Integrity. Trends in Cognitive Sciences 23, 938–951. 10.1016/j.tics.2019.08.003.

39. Thieleking, R., Medawar, E., Disch, L., and Witte, A.V. (2020). art.pics Database: An Open Access Database for Art Stimuli for Experimental Research. Frontiers in Psychology 11. 10.3389/fpsyg.2020.576580.

40. Tian, Y., Margulies, D.S., Breakspear, M., and Zalesky, A. (2020). Topographic organization of the human subcortex unveiled with functional connectivity gradients. Nat Neurosci 23, 1421–1432. 10.1038/s41593-020-00711-6.

41. Adcock, R.A., Thangavel, A., Whitfield-Gabrieli, S., Knutson, B., and Gabrieli, J.D.E. (2006). Reward-Motivated Learning: Mesolimbic Activation Precedes Memory Formation. Neuron 50, 507–517. 10.1016/j.neuron.2006.03.036.

42. Lisman, J.E., and Grace, A.A. (2005). The Hippocampal-VTA Loop: Controlling the Entry of Information into Long-Term Memory. Neuron 46, 703–713. 10.1016/j.neuron.2005.05.002.

43. Rolls, E.T. (2019). Chapter 2 - The cingulate cortex and limbic systems for action, emotion, and memory. In Handbook of Clinical Neurology Cingulate Cortex., B. A. Vogt, ed. (Elsevier), pp. 23–37. 10.1016/B978-0-444-64196-0.00002-9.

44. Egecioglu, E., Skibicka, K.P., Hansson, C., Alvarez-Crespo, M., Friberg, P.A., Jerlhag, E., Engel, J.A., and Dickson, S.L. (2011). Hedonic and incentive signals for body weight control. Rev Endocr Metab Disord 12, 141–151. 10.1007/s11154-011-9166-4.

45. Malik, S., McGlone, F., Bedrossian, D., and Dagher, A. (2008). Ghrelin Modulates Brain Activity in Areas that Control Appetitive Behavior. Cell Metabolism 7, 400–409. 10.1016/j.cmet.2008.03.007.

46. Carlini, V.P., Ghersi, M., Schiöth, H.B., and de Barioglio, S.R. (2010). Ghrelin and memory: Differential effects on acquisition and retrieval. Peptides 31, 1190–1193. 10.1016/j.peptides.2010.02.021.

47. Vartanian, M., Endres, K.J., Lee, Y.T., Friedrich, S., Meemken, M.-T., Schamarek, I., Rohde-Zimmermann, K., Schürfeld, R., Eisenberg, L., Hilbert, A., et al. (2025). Investigating the impact of microbiome-changing interventions on food decision-making: MIFOOD study protocol. BMC Nutr 11, 8. 10.1186/s40795-024-00971-6.

48. Tsaban, G., Yaskolka Meir, A., Zelicha, H., Rinott, E., Kaplan, A., Shalev, A., Katz, A., Brikner, D., Blüher, M., Ceglarek, U., et al. (2022). Diet-induced Fasting Ghrelin Elevation Reflects the Recovery of Insulin Sensitivity and Visceral Adiposity Regression. J Clin Endocrinol Metab 107, 336–345. 10.1210/clinem/dgab681.

49. Davis, C., Mudd, J., and Hawkins, M. (2014). Neuroprotective effects of leptin in the context of obesity and metabolic disorders. Neurobiology of Disease 72, 61–71. 10.1016/j.nbd.2014.04.012.

50. Tang, H., Donahoo, W.T., DeKosky, S.T., Lee, Y.A., Kotecha, P., Svensson, M., Bian, J., and Guo, J. (2025). GLP-1RA and SGLT2i Medications for Type 2 Diabetes and Alzheimer Disease and Related Dementias. JAMA Neurol 82, 439–449. 10.1001/jamaneurol.2025.0353.

51. Bae, J.B., Lee, S., Jung, W., Park, S., Kim, W., Oh, H., Han, J.W., Kim, G.E., Kim, J.S., Kim, J.H., et al. (2020). Identification of Alzheimer’s disease using a convolutional neural network model based on T1-weighted magnetic resonance imaging. Scientific reports 10, 22252. 10.1038/s41598-020-79243-9.

52. Farr, O.M., Li, C.R., and Mantzoros, C.S. (2016). Central Nervous System Regulation of Eating: Insights from Human Brain Imaging. Metabolism 65, 699–713. 10.1016/j.metabol.2016.02.002.

53. Gerges, N.Z., Aleisa, A.M., and Alkadhi, K.A. (2003). Impaired long-term potentiation in obese zucker rats: possible involvement of presynaptic mechanism. Neuroscience 120, 535–539. 10.1016/s0306-4522(03)00297-5.

54. Li, Z., and Liu, J. (2024). Thyroid dysfunction and Alzheimer’s disease, a vicious circle. Front. Endocrinol. 15. 10.3389/fendo.2024.1354372.

55. Ye, J., Huang, Z., Liang, C., Yun, Z., Huang, L., Liu, Y., and Luo, Z. (2024). Thyroid dysfunction and risk of different types of dementia: A systematic review and meta-analysis. Medicine (Baltimore) 103, e39394. 10.1097/MD.0000000000039394.

56. Klein-Flügge, M.C., Bongioanni, A., and Rushworth, M.F.S. (2022). Medial and orbital frontal cortex in decision-making and flexible behavior. Neuron 110, 2743–2770. 10.1016/j.neuron.2022.05.022.

57. Wittmann, B.C., Schott, B.H., Guderian, S., Frey, J.U., Heinze, H.-J., and Düzel, E. (2005). Reward-Related fMRI Activation of Dopaminergic Midbrain Is Associated with Enhanced Hippocampus-Dependent Long-Term Memory Formation. Neuron 45, 459–467. 10.1016/j.neuron.2005.01.010.

58. Brebner, L.S., Ziminski, J.J., Margetts-Smith, G., Sieburg, M.C., Hall, C.N., Heintz, T.G., Lagnado, L., Hirrlinger, J., Crombag, H.S., and Koya, E. (2020). Extinction of cue-evoked food-seeking recruits a GABAergic interneuron ensemble in the dorsal medial prefrontal cortex of mice. European Journal of Neuroscience 52, 3723–3737. 10.1111/ejn.14754.

59. Struik, R.F., Marchant, N.J., De Haan, R., Terra, H., Van Mourik, Y., Schetters, D., Carr, M.R., Van Der Roest, M., Heistek, T.S., and De Vries, T.J. (2019). Dorsomedial prefrontal cortex neurons encode nicotine-cue associations. Neuropsychopharmacol. 44, 2011–2021. 10.1038/s41386-019-0449-x.

60. Xia, H., Wu, Q., Shields, G.S., Nie, H., Hu, X., Liu, S., Zhou, Z., Chen, H., and Yang, Y. (2024). Neural activity and connectivity are related to food preference changes induced by food go/no-go training. Neuropsychologia 201, 108919. 10.1016/j.neuropsychologia.2024.108919.

61. Holroyd, C.B., Ribas-Fernandes, J.J.F., Shahnazian, D., Silvetti, M., and Verguts, T. (2018). Human midcingulate cortex encodes distributed representations of task progress. Proceedings of the National Academy of Sciences 115, 6398–6403. 10.1073/pnas.1803650115.

62. Kolling, N., Wittmann, M.K., Behrens, T.E.J., Boorman, E.D., Mars, R.B., and Rushworth, M.F.S. (2016). Value, search, persistence and model updating in anterior cingulate cortex. Nat Neurosci 19, 1280–1285. 10.1038/nn.4382.

63. Amemori, K., and Graybiel, A.M. (2012). Localized microstimulation of primate pregenual cingulate cortex induces negative decision-making. Nat Neurosci 15, 776–785. 10.1038/nn.3088.

64. Kringelbach, M.L., and Rolls, E.T. (2004). The functional neuroanatomy of the human orbitofrontal cortex: evidence from neuroimaging and neuropsychology. Prog Neurobiol 72, 341–372. 10.1016/j.pneurobio.2004.03.006.

65. Seabrook, L.T., and Borgland, S.L. (2020). The orbitofrontal cortex, food intake and obesity. J Psychiatry Neurosci 45, 304–312. 10.1503/jpn.190163.

66. Arnoriaga-Rodríguez, M., Mayneris-Perxachs, J., Burokas, A., Contreras-Rodríguez, O., Blasco, G., Coll, C., Biarnés, C., Miranda-Olivos, R., Latorre, J., Moreno-Navarrete, J.-M., et al. (2020). Obesity Impairs Short-Term and Working Memory through Gut Microbial Metabolism of Aromatic Amino Acids. Cell Metab 32, 548–560.e7. 10.1016/j.cmet.2020.09.002.

67. Blechert, J., Lender, A., Polk, S., Busch, N.A., and Ohla, K. (2019). Food-Pics_Extended—An Image Database for Experimental Research on Eating and Appetite: Additional Images, Normative Ratings and an Updated Review. Frontiers in Psychology 10.

68. Medawar, E., Thieleking, R., and Witte, A.V. (2022). Dietary Fiber and WHO Food Categories Extension for the Food-Pics_Supplementary base. Front. Psychol. 13. 10.3389/fpsyg.2022.818471.

69. Berding, K., Long-Smith, C.M., Carbia, C., Bastiaanssen, T.F.S., Van De Wouw, M., Wiley, N., Strain, C.R., Fouhy, F., Stanton, C., Cryan, J.F., et al. (2021). A specific dietary fibre supplementation improves cognitive performance—an exploratory randomised, placebo-controlled, crossover study. Psychopharmacology 238, 149–163. 10.1007/s00213-020-05665-y.

70. Freijy, T.M., Cribb, L., Oliver, G., Metri, N.-J., Opie, R.S., Jacka, F.N., Hawrelak, J.A., Rucklidge, J.J., Ng, C.H., and Sarris, J. (2024). The impact of a prebiotic-rich diet and/or probiotic supplements on human cognition: Secondary outcomes from the ‘Gut Feelings’ randomised controlled trial. Nutritional Neuroscience, 1–11. 10.1080/1028415X.2024.2425570.

71. Esteban, O., Markiewicz, C.J., Blair, R.W., Moodie, C.A., Isik, A.I., Erramuzpe, A., Kent, J.D., Goncalves, M., DuPre, E., Snyder, M., et al. (2019). fMRIPrep: a robust preprocessing pipeline for functional MRI. Nat Methods 16, 111–116. 10.1038/s41592-018-0235-4.

72. Esteban, O., Birman, D., Schaer, M., Koyejo, O.O., Poldrack, R.A., and Gorgolewski, K.J. (2017). MRIQC: Advancing the automatic prediction of image quality in MRI from unseen sites. PLOS ONE 12, e0184661. 10.1371/journal.pone.0184661.

73. Smith, S.M., Jenkinson, M., Woolrich, M.W., Beckmann, C.F., Behrens, T.E.J., Johansen-Berg, H., Bannister, P.R., De Luca, M., Drobnjak, I., Flitney, D.E., et al. (2004). Advances in functional and structural MR image analysis and implementation as FSL. NeuroImage 23, S208–S219. 10.1016/j.neuroimage.2004.07.051.

74. Jensen, D.E.A., Ebmeier, K.P., Suri, S., Rushworth, M.F., and Klein-Flügge, M.C. (2024). Nuclei-specific hypothalamus networks predict a dimensional marker of stress in humans. Nature Communications. 10.1038/s41467-024-46275-y.

75. Klein-Flügge, M., Jensen, D., Tagaki, Y., Priestley, L., Verhagen, L., Smith, S., and Rushworth, M. (2022). Relationship between nuclei-specific amygdala connectivity and mental health dimensions in humans. Nat Hum Beh.

76. Guillaume, B., Hua, X., Thompson, P.M., Waldorp, L., and Nichols, T.E. (2014). Fast and accurate modelling of longitudinal and repeated measures neuroimaging data. NeuroImage 94, 287–302. 10.1016/j.neuroimage.2014.03.029.

