## Supplementary for "Neural Correlates of Human Food Memory link to Microbial, Homeostatic, and Hedonic Signals: Evidence from a Prebiotic Randomized Clinical Trial"

#### Title

### 1. Participants demographics

***Supplementary Table 1: Participants' demographics at the first study visit and baseline and follow-up prebiotic and placebo groups.*** *Presented are estimated means with +- standard deviations and the range (min, max).*

| | *first study visit* | *prebiotic group* | | *placebo group* | |
|---|---|---|---|---|---|
| | | **BL** | **FU** | **BL** | **FU** |
| *Sample (N)* | 55 | 52 | 45 | 50 | 47 |
| *Age* | 28.36 ± 6.54 (19.00, 45.00) | 28.58 ± 6.38 (19.00, 45.00) | 28.29 ± 6.18 (19.00, 45.00) | 28.22 ± 6.31 (19.00, 45.00) | 28.45 ± 6.11 (19.00, 45.00) |
| *Sex* | M: 36, F: 19 | M: 36, F: 16 | M: 33, F: 12 | M: 34, F: 16 | M: 34, F: 13 |
| *SES* | 14.63 ± 2.94 (5.10, 19.20) | 14.56 ± 2.99 (5.10, 19.20) | 14.60 ± 3.10 (5.10, 19.20) | 14.68 ± 3.00 (5.10, 19.20) | 14.61 ± 3.05 (5.10, 19.20) |
| *BMI [kg/m$^2$]* | 27.24 ± 1.49 (25.00, 30.00) | 27.18 ± 1.48 (24.50, 30.20) | 27.30 ± 1.59 (24.50, 30.60) | 27.32 ± 1.59 (25.00, 31.20) | 27.24 ± 1.62 (24.90, 31.30) |
| *WHR [waist/height]* | 0.82 ± 0.06 (0.70, 0.95) | 0.82 ± 0.06 (0.70, 0.94) | 0.81 ± 0.06 (0.69, 0.97) | 0.82 ± 0.06 (0.69, 0.98) | 0.81 ± 0.06 (0.71, 0.98) |
| *FM [%]* | 27.15 ± 6.69 (7.59, 39.76) | 26.43 ± 6.59 (7.59, 38.48) | 26.24 ± 6.34 (10.57, 38.97) | 26.89 ± 6.77 (9.53, 41.58) | 26.03 ± 6.57 (7.76, 38.86) |
| *Diastolic blood pressure* | 77.65 ± 8.48 (62.67, 98.67) | 76.61 ± 8.74 (61.33, 98.67) | 76.84 ± 8.64 (62.33, 95.00) | 76.99 ± 8.50 (61.33, 98.00) | 76.35 ± 8.52 (59.67, 92.00) |
| *Systolic blood pressure* | 128.48 ± 11.26 (105.33, 152.67) | 127.68 ± 11.52 (105.33, 146.33) | 126.32 ± 10.72 (107.33, 144.00) | 127.81 ± 10.83 (106.67, 152.67) | 126.48 ± 10.76 (106.67, 149.00) |
| *HDL* | 49.70 ± 10.93 (29.00, 84.00) | 50.02 ± 11.20 (25.00, 77.00) | 49.89 ± 13.11 (27.00, 79.00) | 49.83 ± 11.04 (29.00, 84.00) | 49.27 ± 11.56 (26.00, 77.00) |
| *LDL* | 99.54 ± 25.96 (35.00, 160.00) | 97.67 ± 24.64 (35.00, 160.00) | 102.20 ± 33.95 (44.00, 236.00) | 99.40 ± 28.19 (46.00, 154.00) | 93.49 ± 24.47 (53.00, 143.00) |
| *Energy intake [kcal]* | 1599.15 ± 511.65 (500.87, 2727.30) | 1618.23 ± 544.91 (500.87, 2777.99) | 1585.36 ± 448.04 (494.89, 2728.47) | 1636.53 ± 443.02 (627.58, 2496.09) | 1527.69 ± 485.18 (682.63, 2608.66) |

*Abbreviations: BL - baseline, FU - follow-up, SES - sociodemographic status, BMI - body-mass-index, WHR - waist-to-hip ratio, FM - fat mass, HDL - high-density lipoprotein, LDL - low-density lipoprotein.*

### 2. Descriptive statistics for memory accuracy

***Supplementary Table 2: Means +- standard deviations (SD), range of encoding and retrieval memory accuracy estimates.***

| | **Encoding** | | **Retrieval** | |
|---|---|---|---|---|
| | **Food** | **Art** | **Food** | **Art** |
| **Memory accuracy** (mean+- SD, range; in %) | 0.81+-0.10 (0.53, 1.0) | 0.70+-0.079 (0.51, 0.93) | 0.81+-0.07 (0.57, 0.94) | 0.48+-0.04 (0.36, 0.59) |
| **Hit accuracy rate** (mean+- SD, range; in %) | 0.81+-0.10 (0.53, 1.0) | 0.70+-0.08 (0.51, 0.93) | 0.30+-0.04 (0.2, 0.38) | 0.26+-0.03 (0.19, 0.35) |
| **Miss accuracy rate** (mean+- SD, range; in %) | 0.30+-0.17 (0.08, 1.0) | 0 * no misses | 0.11+-0.06 (0.03, 0.37) | 0.18+-0.07 (0.08, 0.34) |
| **False alarm accuracy rate** (mean+- SD, range; in %) | .. | .. | 0.07+-0.03 (0.01, 0.18) | 0.11+-0.03 (0.03, 0.15) |
| **Correct rejection accuracy rate** (mean+- SD, range; in %) | .. | .. | 0.51+-0.06 (0.26, 0.60) | 0.43+-0.07 (0.27, 0.54) |

#### 3. Preregistered Hypotheses

**Supplementary *Table 3: Preregistered Hypotheses.***

| **Label in Manuscript** | **Label in Preregistration** | **Hypothesis** | **TFCE-FWE** | **Voxel-wise-FWE** | **TFCE** | **Voxel-wise** |
|---|---|---|---|---|---|---|
| **Memory *wanting** | | | | | | |
| H1_m | H3_m | Food compared to art reward-enhancement of recognition memory accuracy is associated with differences in BOLD activation of reward ROIs during encoding. | ✓ | ✓ | ✓ | ✓ |
| H1_e | H8_e | Food compared to art reward-enhancement of memory accuracy is associated with differences in BOLD activation of reward ROIs during retrieval. | ✓ | ✓ | ✓ | ✓ |
| **Food memory** | | | | | | |
| H2_m | H2_m | Food compared to art recognition memory accuracy is associated with differences in BOLD activation during encoding. | ✓ | ✓ | ✓ | ✓ |
| H3_m | H1_m | Food compared to art elicits different recognition memory accuracy (defined as correct recognition of 'old' stimuli or correct identification of new stimuli) which is associated with differences in BOLD activation of the hippocampus during retrieval. | ☒ (but ✓ without correction with sex) | ☒ | ✓ | ✓ |
| H2_e | H7_e | Food compared to art recognition/retrieval memory accuracy is associated with differences in BOLD activation of ROIs other than the hippocampus. | ☒ (but ✓ without correction with sex) | ☒ | ✓ | ✓ |
| **Intervention** | | | | | | |
| H3_e | H10_e | Neural correlates memory retrieval accuracy for food versus art is increased by prebiotic intervention by showing an increased hippocampal activation. Furthermore, the interventional effect of a prebiotic diet influenced the microbiome, inflammatory markers and shows memory-related changes in the BOLD signal. | ✓ (but reduced activation) | ✓ | ✓ | ✓ |
| H4_e | H14_e | Memory accuracy during encoding of wanted stimuli (and food versus art) is increased after prebiotic intervention and shows differences in BOLD activation in the hippocampus. | ☒ | ☒ | ✓ | ✓ |
| H5_e | H15_e | Based on findings from Medawar et al., (2023), memory retrieval accuracy of wanted stimuli (and food versus art) is increased after prebiotic intervention and shows differences in BOLD activation in reward network ROIs. | ☒ | ☒ | ✓ | ✓ |
| **Microbiota/blood** | | | **Outcome** | | | |
| H4_m | H6_m | The BOLD activation of the hippocampus during retrieval and/or encoding is associated with food versus art memory accuracy and differences in microbiota (i.e., diversity and abundance). | ✓ (parahippocampal activations but not related to microbiota) | | | |
| H6_e | H12_e | Memory accuracy is increased by prebiotic intervention, dependent on microbiome | ☒ | | | |

| | | | |
|---|---|---|---|
| | | composition and BOLD hippocampal activation during encoding and retrieval. | |
| H7_e | H11_e | The BOLD activation of the hippocampus during retrieval and/or encoding is associated with intervention-induced (prebiotics) changes in small chain fatty acids (SCFAs, i.e. fecal acetate, butyrate, propionate) and inflammatory markers (i.e., IL-6, high-reactive C-reactive protein (hsCRP), tumor necrosis factor α (TNF-α), and trimethylamine N-oxide (TMAO)). | ✓ |
| H8_e | H13_e | Memory accuracy for food versus art is increased by prebiotic intervention, dependent on microbiome composition and BOLD hippocampal activation at encoding and retrieval. | ✓ |
| **Caloric bias** | | | |
| H5_m | H4_m | The calorie bias of the food memory is associated with differences in BOLD activation at encoding and retrieval (independent of reward expectation). | ☒ |
| H9_e | H9_e | BOLD activations towards memory accuracy during encoding are reward-enhanced but shows decreased activations for high-caloric food after a prebiotic intervention. | ☒ |

#### 4. Average neural activations during successful versus non-successful memory encoding and retrieval

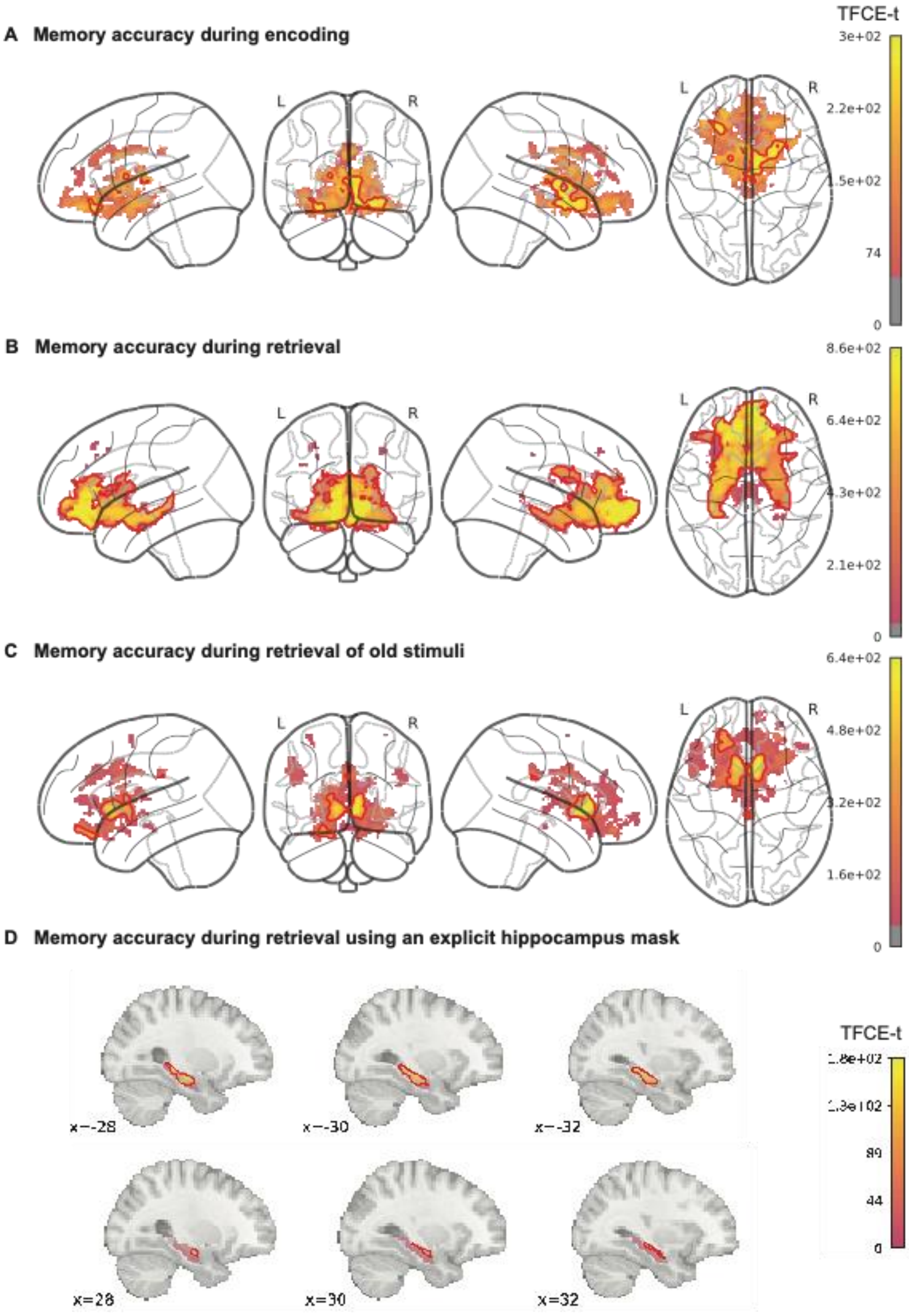

**Supplementary** ***Figure 1: Neural response to food and art stimuli memory encoding and retrieval.***

*(A) Memory accuracy during encoding (correct identification of old images), (B) during retrieval (correct identification of old images and correct rejection of new stimuli), and (C) retrieval of old stimuli (correct identification of old images) elicited neural activations in hedonic and homeostatic brain regions. Correct identification of old stimuli during retrieval showed lower neural activations compared to, but higher than during encoding. (D) Using an explicit hippocampus mask (Tian et al., 2020), memory accuracy during retrieval elicited neural activations in the hippocampus head and body. Statistics were done on voxel-wise blood-oxygen-level-dependent signal using the sandwich estimator toolbox with threshold-free cluster enhancement (TFCE) family-wise-error correction (FWE) of multiple comparisons (main analyses) and covariates of sex and wanting of individual stimuli. Color bars depict parametric TFCE statistic (TFCE-t>50 for visualization purposes) with wild-boot strapped pFWE<0.05 marked in red outline.*

**Supplementary *Table 4: Localization of significant changes in brain activation to visual food and art stimuli during functional MRI memory encoding and retrieval.*** *Neural activations are shown either across groups (prebiotic and placebo, main) or in the interaction of intervention groups (prebiotic, placebo) * time points (baseline, follow-up). Extracted clusters larger than 500 $mm^3$ are indicated in bold. Total size and cluster sizes are given in $mm^{3,}$ and peak locations are given at mm.*

| ***Figure*** | **Model (main/ interaction)** | **total size** | **No** | **size** | **pTFCE +FWE** | **pFWE** | **pFDR** | **Z** | **pUnc** | **x** | **y** | **z** | **Region** |
|---|---|---|---|---|---|---|---|---|---|---|---|---|---|
| *1C* | encoding * wanting modulation (main) | 39008 | 1 | **38904** | 0.001 | 0 | 0.004 | 6.12 | 0.001 | -8 | 30 | 22 | Cingulate Ant L |
| | | | 2 | 104 | 0.004 | 0 | 0.004 | 5.18 | 0.001 | -30 | 14 | 54 | Frontal Mid 2 L |
| *1D* | retrieval * wanting modulation, uncorrected for sex (main) | 4168 | 1 | 312 | 0.029 | 0 | 0.023 | 5.01 | 0.001 | -2 | -28 | 34 | Cingulate Mid L |
| | | | 2 | 176 | 0.034 | 0 | 0.023 | 4.79 | 0.001 | -30 | 24 | -6 | Insula L |
| | | | 3 | 624 | 0.02 | 0 | 0.023 | 4.75 | 0.001 | 52 | 30 | 24 | Frontal Inf Tri R / dmPFC |
| | | | 4 | **3056** | 0.009 | 0.01 | 0.023 | 4.53 | 0.001 | -50 | 32 | 28 | Frontal Inf Tri L/ dmPFC |
| *1E* | food > art encoding *wanting modulation (main) | 32 | 1 | 24 | 0.022 | 0.01 | 0.151 | 4.37 | 0.001 | 2 | 40 | 0 | Cingulate Ant R/ACC |
| | | | 2 | 8 | 0.049 | 0.1 | 0.151 | 3.64 | 0.001 | 4 | 42 | -2 | Cingulate Ant R/ACC |
| *S1A* | encoding (main) | 3632 | 1 | 496 | 0.019 | 0.08 | 0.081 | 4.19 | 0.001 | -24 | 28 | -18 | OFC post L |
| | | | 2 | 72 | 0.04 | 0.13 | 0.081 | 3.96 | 0.001 | -16 | 2 | 10 | Caudate L (WM) |
| | | | 3 | **3040** | 0.017 | 0.15 | 0.081 | 3.91 | 0.001 | 2 | -6 | 4 | Thalamus R (towards amygdala/hippocampal area) |
| | | | 4 | 16 | 0.049 | 0.48 | 0.081 | 3.34 | 0.001 | -4 | 8 | -4 | Caudate L |
| | | | 5 | 8 | 0.05 | 0.52 | 0.081 | 3.29 | 0.001 | -6 | 12 | -2 | Caudate L |
| *S1B* | retrieval (main) | 50128 | 1 | **50128** | 0.001 | 0.01 | 0.012 | 4.76 | 0.001 | 8 | 36 | 8 | Cingulate Ant R |
| *S1C* | retrieval old (main) | 6136 | 1 | **5400** | 0.002 | 0 | 0.031 | 5.3 | 0.001 | 6 | 16 | -2 | Caudate R |
| | | | 2 | 104 | 0.042 | 0 | 0.031 | 5.04 | 0.001 | -2 | -30 | 28 | Cingulate posterior |
| | | | 3 | **632** | 0.013 | 0 | 0.031 | 5.02 | 0.001 | -22 | 32 | -24 | Frontal pole |
| *2A* | food > art encoding (main) | 8768 | 1 | **8448** | 0.015 | 0.18 | 0.068 | 3.83 | 0.001 | 28 | -4 | -6 | Pallidum R |
| | | | 2 | 232 | 0.038 | 0.25 | 0.068 | 3.68 | 0.001 | 16 | 8 | 20 | Caudate R |
| | | | 3 | 88 | 0.048 | 0.57 | 0.068 | 3.21 | 0.001 | 16 | -6 | 26 | Caudate R |

| | | | | | | | | | | | | | |
|---|---|---|---|---|---|---|---|---|---|---|---|---|---|
| *2B* | food > art retrieval, uncorrected for sex (main) | 824 | 1 | **800** | 0.029 | 0.03 | 0.172 | 4.2 | 0.001 | -18 | -6 | -16 | Amygdala L |
| | | | 2 | 24 | 0.048 | 0.08 | 0.172 | 3.96 | 0.001 | 18 | -8 | -16 | Amygdala R |
| *S2A* | food retrieval (main) | 32496 | 1 | **32320** | 0.001 | 0.01 | 0.025 | 4.51 | 0.001 | 24 | -8 | -14 | Amygdala R |
| | | | 2 | 176 | 0.037 | 0.19 | 0.031 | 3.51 | 0.002 | -26 | -4 | 10 | Putamen L |
| *S2B* | art retrieval (main) | 20064 | 1 | **19200** | 0.002 | 0 | 0.045 | 5.31 | 0.001 | 4 | 16 | -4 | Olfactory R |
| | | | 2 | **640** | 0.029 | 0.03 | 0.045 | 4.36 | 0.001 | 22 | 6 | 26 | WM (near Caudate R) |
| | | | 3 | 16 | 0.039 | 0.05 | 0.045 | 4.14 | 0.001 | 26 | 12 | 22 | WM |
| | | | 4 | 192 | 0.036 | 0.1 | 0.045 | 3.94 | 0.001 | -22 | 4 | -20 | Temporal Pole Sup L |
| | | | 5 | 16 | 0.05 | 0.78 | 0.055 | 2.76 | 0.003 | -20 | 22 | 10 | WM |
| - | food retrieval old (main) | 5384 | 1 | **2568** | 0.004 | 0.002 | 0.047 | 5.775 | 0.001 | -10 | 16 | 0 | Caudate L |
| | | | 2 | **2192** | 0.012 | 0.036 | 0.047 | 4.545 | 0.001 | 8 | 16 | 0 | Caudate R |
| | | | 3 | **624** | 0.027 | 0.14 | 0.047 | 3.977 | 0.001 | -14 | 2 | -8 | Pallidum L |
| *3B (top)* | food > art encoding (interaction) | 1056 | 1 | **864** | 0.033 | 0.12 | 0.182 | 3.83 | 0.001 | 4 | 36 | -8 | Cingulate Ant R |
| | | | 2 | 152 | 0.043 | 0.4 | 0.182 | 3.33 | 0.001 | -2 | 32 | 2 | Cingulate Ant L, Paracingulate |
| | | | 3 | 40 | 0.05 | 0.57 | 0.182 | 3.15 | 0.001 | 0 | 56 | -4 | Frontal Med Orb L |
| *3B (middle)* | food > art retrieval (interaction) | 4152 | 1 | **1240** | 0.011 | 0.01 | 0.054 | 4.65 | 0.001 | -44 | 16 | 42 | Frontal Mid 2 L |
| | | | 2 | **736** | 0.009 | 0.01 | 0.054 | 4.63 | 0.001 | 0 | 18 | 26 | Cingulate Ant L |
| | | | 3 | 336 | 0.034 | 0.06 | 0.054 | 4.16 | 0.001 | 42 | 22 | 52 | Frontal Mid 2 R |
| | | | 4 | **768** | 0.03 | 0.14 | 0.054 | 3.84 | 0.001 | -8 | 44 | 0 | Cingulate Ant L |
| | | | 5 | **640** | 0.031 | 0.2 | 0.054 | 3.7 | 0.001 | -2 | 38 | 10 | Cingulate Ant L |
| | | | 6 | 432 | 0.041 | 0.35 | 0.054 | 3.45 | 0.001 | -8 | 44 | 12 | Cingulate Ant L |
| *3B (bottom)* | food > art retrieval old (interaction) | 33816 | 1 | **24440** | 0.009 | 0.06 | 0.064 | 4.18 | 0.001 | 16 | 10 | 14 | Caudate R (incl. VTA, hippocampus, thalamus) |
| | | | 2 | **9376** | 0.014 | 0.12 | 0.064 | 3.94 | 0.001 | 4 | 10 | 40 | Cingulate Mid R (incl ACC) |
| *4A* | retrieval, 1st visit (main) | 6376 | 1 | **2352** | 0.012 | 0.05 | 0.076 | 4.15 | 0.001 | -10 | 14 | -12 | Caudate L, Subcallosal cortex, NAc |
| | | | 2 | **3096** | 0.019 | 0.18 | 0.081 | 3.71 | 0.002 | 6 | 62 | -2 | Frontal Med Orb R |
| | | | 3 | 184 | 0.037 | 0.2 | 0.081 | 3.67 | 0.002 | -12 | -4 | -20 | ParaHippocampal L, Amygdala L |
| | | | 4 | 616 | 0.036 | 0.35 | 0.081 | 3.4 | 0.002 | -12 | 48 | -8 | Frontal Med Orb L |
| | | | 5 | 64 | 0.044 | 0.37 | 0.076 | 3.37 | 0.001 | -8 | 24 | -2 | near Accumbens L |
| | | | 6 | 40 | 0.046 | 0.52 | 0.076 | 3.17 | 0.001 | -12 | 38 | -20 | Rectus L |
| | | | 7 | 16 | 0.048 | 0.55 | 0.081 | 3.14 | 0.002 | -16 | -6 | -26 | ParaHippocampal L |
| | | | 8 | 8 | 0.048 | 0.71 | 0.098 | 2.93 | 0.004 | -12 | 0 | -12 | near Accumbens L |
| - | art retrieval, 1st visit (main) | 1344 | 1 | **1264** | 0.018 | 0.03 | 0.132 | 4.19 | 0.001 | 14 | 32 | -4 | Subcallosal cortex, NAc |
| | | | 2 | 80 | 0.047 | 0.24 | 0.132 | 3.5 | 0.001 | -10 | 14 | -10 | Caudate L, NAc |

*Abbreviations: L - left, R - right, No - cluster number, NAc - nucleus accumbens, Med - medial, Orb - orbital, VTA - ventral tegmentum area, Mid - middle, ACC - anterior cingulate cortex, Ant – anterior, WM - white matter.*

#### 5. Neural activations within the hippocampus

**Supplementary *Table 5: Localization of significant changes in hippocampal brain activation to visual food and art stimuli during functional MRI memory encoding and retrieval.*** *Neural activations are shown either across groups (prebiotic and placebo, main) or in the interaction of intervention groups (prebiotic, placebo) * time points (baseline, follow-up) and restricted to an explicit hippocampus mask (Tian et al., 2020). Extracted clusters larger than 500 mm$^3$ are indicated in bold. Total size and cluster sizes are given in mm$^{3,}$ and peak locations are given at mm.*

| Figure | Model - using explicit hippocampus masks (main/interaction) | total size | cluster | size | pTFCE +FWE | pFWE | pFDR | Z | pUnc | x | y | z | Subregion |
|---|---|---|---|---|---|---|---|---|---|---|---|---|---|
| *S1D* | retrieval (main) | 3216 | 1 | **872** | 0.011 | 0.008 | 0.013 | 3.859 | 0.001 | 28 | -10 | -16 | R head |
| | | | 2 | **2344** | 0.002 | 0.008 | 0.013 | 3.853 | 0.001 | -32 | -30 | -6 | L body |
| *S3A* | food > art encoding (interaction) | 640 | 1 | **640** | 0.015 | 0.035 | 0.059 | 3.533 | 0.001 | 24 | -22 | -14 | R body |
| *S3B* | food > art retrieval old (interaction) | 192 | 1 | 64 | 0.04 | 0.069 | 0.147 | 3.316 | 0.002 | 32 | -30 | -6 | R body |
| | | | 2 | 128 | 0.042 | 0.16 | 0.147 | 3.004 | 0.001 | 28 | -18 | -12 | R body |

*Abbreviations: L - left, R - right.*

#### 6. Neural activations during memory retrieval for food and art stimuli (individually)

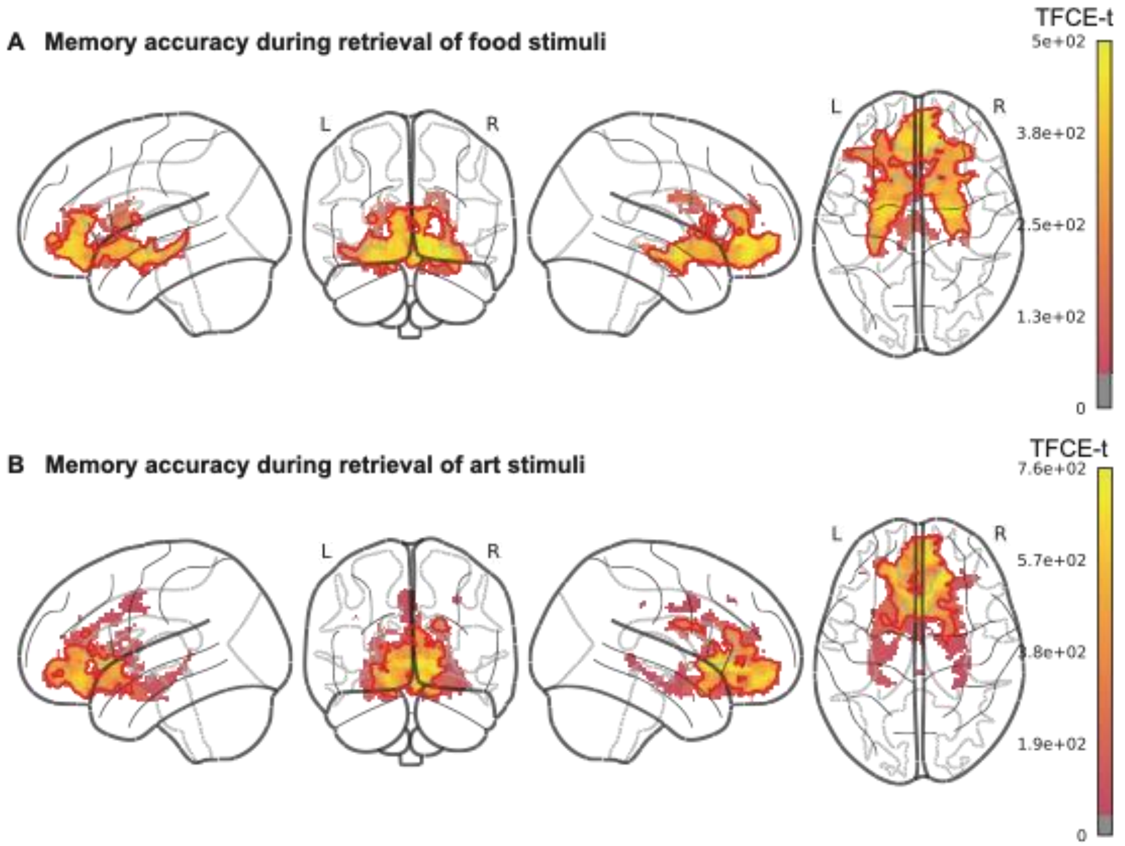

***Supplementary Figure 2: Neural response during memory retrieval for food (A) and art (B) stimuli elicited neural activations in hedonic brain regions.*** *Statistics were done on voxel-wise blood-oxygen-level-dependent signal using the sandwich estimator toolbox with threshold-free cluster enhancement (TFCE) family-wise-error correction (FWE) of multiple comparisons (main analyses) and covariates of sex and wanting of individual stimuli. Color bars depict parametric TFCE statistic (TFCE-t>50 for visualization purposes) with wild-boot strapped* $p_{FWE}$*<0.05 marked in red outline.*

#### 7. Linear Mixed Effect Model Results of Memory Accuracy related to prebiotic intervention

**Supplementary** ***Table 6: Linear mixed effect model results for changes in memory accuracy for post-prebiotic intervention compared to placebo***. *Significant Formula: variable ~ intervention * timepoint*

*+ (1+(timepoint+intervention)|subject)+ age + sex. Only significant group*time interaction effects are shown here; the extended Table is shown in* Supplementary *Table 11.*

| **Variable** | **Observations** | **Groups** | **Effect** | **Estimate** | **SE** | **T value** | **p value** |
|---|---|---|---|---|---|---|---|
| *PSimFood* | 175 | 55 | (Intercept) | 0.78 | 0.03 | 23.43 | |
| | | | timepoint FU | 0.04 | 0.02 | 1.62 | |
| | | | interventionprebiotic | 0.04 | 0.02 | 2.09 | |
| | | | sex (male) | -0.1 | 0.04 | -2.72 | |
| | | | timepoint FU:interventionprebiotic | -0.08 | 0.03 | -2.64 | **0.009** |
| *RetFoodType2* | 175 | 55 | (Intercept) | 10.84 | 0.72 | 15.02 | |
| | | | timepoint FU | -0.22 | 0.97 | -0.23 | |
| | | | interventionprebiotic | 0.28 | 0.88 | 0.32 | |
| | | | sex (male) | -0.61 | 0.56 | -1.09 | |
| | | | timepoint FU:interventionprebiotic | -0.57 | 1.05 | -0.54 | **0.003** |
| *EncFoodCal1* | 175 | 55 | (Intercept) | 5.14 | 0.31 | 16.61 | |
| | | | timepoint FU | 0.31 | 0.34 | 0.92 | |
| | | | interventionprebiotic | 0.33 | 0.33 | 0.99 | |
| | | | sex (male) | 0.1 | 0.3 | 0.33 | |
| | | | timepoint FU:interventionprebiotic | -1.19 | 0.46 | -2.6 | **0.009** |

*Abbreviations: FU - follow-up, PSimFood - Percentage of similar food stimuli, RetFoodType2 - Retrieval of Food stimuli of type 2 (fruits), EncFoodCal1 - Encoding of food stimuli with caloric content category 1.*

### 8. Neural activations within the hippocampus related to prebiotic intervention

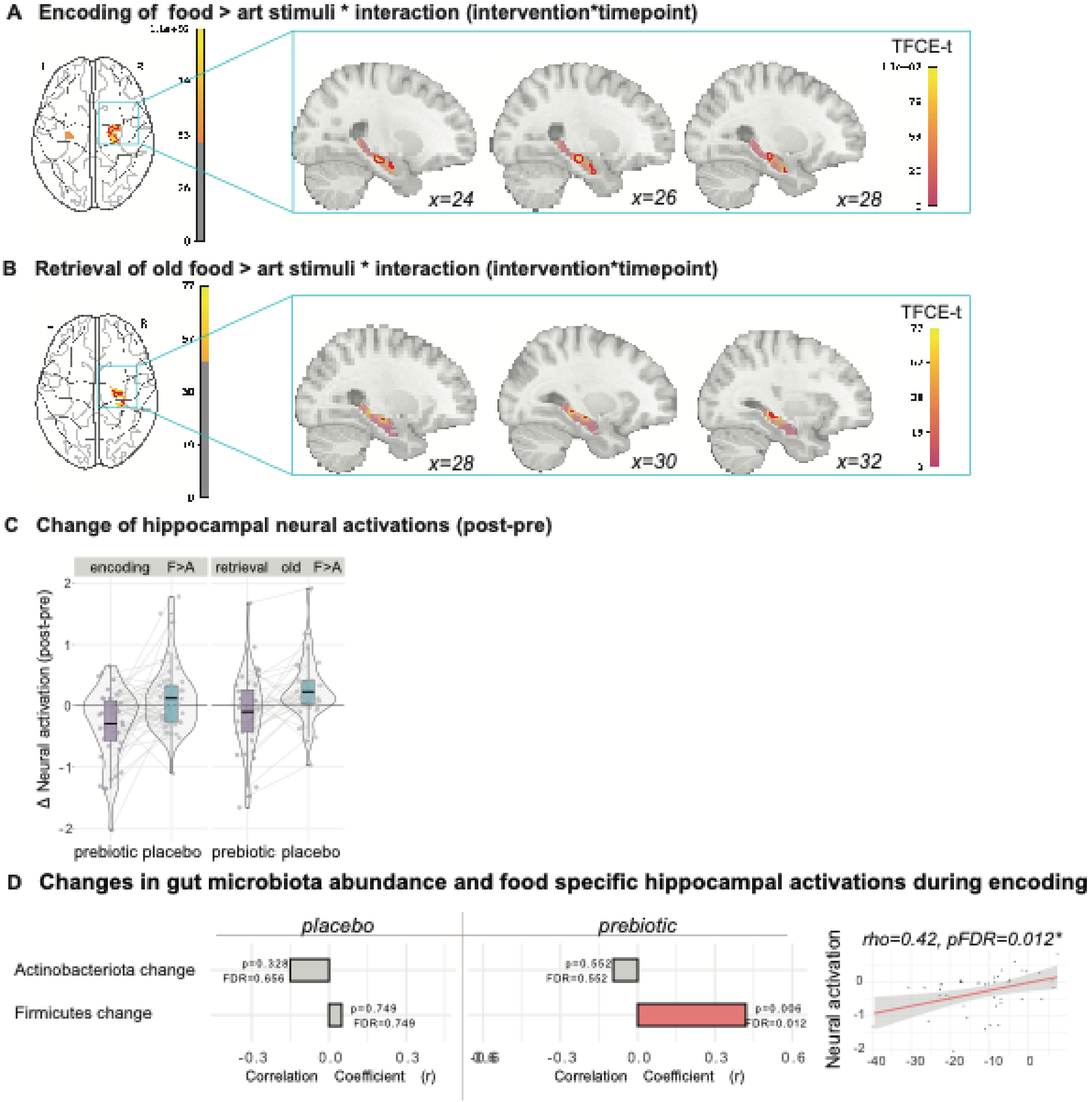

***Supplementary Figure 3: Effects of prebiotic intervention on food memory encoding and retrieval using an explicit hippocampus mask.*** *(A) Food (F) compared to art (A) memory encoding, and (B) elicited neural response in the hippocampus body. Statistics were done on voxel-wise blood-oxygen-level-dependent signal using the sandwich estimator toolbox with threshold-free cluster enhancement (TFCE) family wise-error correction (FWE) of multiple comparisons. Colour bars depict parametric TFCE statistic with wild-boot strapped $p_{FWE}$<0.05 marked in red outline (upper right panel) and as an enlargement (lower right panel). (C) The difference (delta) between baseline and follow-up (post-pre) for neural activations in extracted clusters of the hippocampus are presented for prebiotic and placebo groups. The brain activation responses are shown to decrease in the prebiotic group compared to placebo. (D) Correlation between the change of neural activations (post-to-pre intervention difference – delta), extracted from 2$^{nd}$ level analysis (interaction effects) using an explicit hippocampal mask (Tian et. al., 2020), and the change of gut microbiome diversity. Data is separated by intervention groups showing correlations in the placebo (left) and prebiotic (right) groups. Red bars indicate direct*

*correlations, blue bars indicate inverse correlations and grey bars are non-significant correlations using p<0.05 before FDR correction. Scatterplots on the right show sample distributions.*

### 9. Linear Mixed Effect Model Results of microbiota and blood outcomes related to prebiotic intervention

**Supplementary** ***Table 7: Linear mixed effect model results for changes in fecal and blood markers for post-prebiotic intervention compared to placebo.*** *Significant Formula: variable ~ intervention * timepoint + (1+(timepoint+intervention)|subject)+ age + sex. Only significant group*time interaction effects are shown here which were also shown in Figure 5 (main manuscript); the extended Table is shown in* Supplementary *Table 12.*

| Category | Variable | Obser-vations | Groups | Effect | Estimate | SE | T value | p value |
|---|---|---|---|---|---|---|---|---|
| gut microbiome diversity | Evenness | 192 | 55 | (Intercept) | 0.61 | 0 | 485.78 | |
| | | | | timepoint FU | 0 | 0 | 0.51 | |
| | | | | intervention prebiotic | 0 | 0 | 2.58 | |
| | | | | sex (male) | 0 | 0 | -0.42 | |
| | | | | timepoint FU:intervention prebiotic | -0.01 | 0 | -5.42 | **<0.001** |
| | Richness | 192 | 55 | (Intercept) | 451.34 | 14.35 | 31.45 | |
| | | | | timepoint FU | -3.08 | 12.16 | -0.25 | |
| | | | | intervention prebiotic | -13.3 | 11.89 | -1.12 | |
| | | | | sex (male) | -7.38 | 15.43 | -0.48 | |
| | | | | timepoint FU:intervention prebiotic | -50.55 | 16.35 | -3.09 | **0.002** |
| | shannon effective | 192 | 55 | (Intercept) | 211.99 | 6.37 | 33.26 | |
| | | | | timepoint FU | -1.09 | 5.42 | -0.2 | |
| | | | | intervention prebiotic | -0.44 | 5.23 | -0.08 | |
| | | | | sex (male) | -2.12 | 6.65 | -0.32 | |
| | | | | timepoint FU:intervention prebiotic | -34.63 | 7.39 | -4.69 | **<0.001** |
| gut microbiome abundance (phylum) | Actino-bacteriota | 192 | 55 | (Intercept) | 13.25 | 1.52 | 8.73 | |
| | | | | timepoint FU | -0.73 | 1.09 | -0.67 | |
| | | | | intervention prebiotic | -1.08 | 1.16 | -0.92 | |
| | | | | sex (male) | 1.37 | 1.7 | 0.81 | |
| | | | | timepoint FU:intervention prebiotic | 12.39 | 1.43 | 8.65 | **<0.001** |
| | Firmicutes | 192 | 55 | (Intercept) | 71.38 | 1.34 | 53.18 | |
| | | | | timepoint FU | 0.45 | 1.26 | 0.36 | |
| | | | | intervention prebiotic | 0.38 | 1.31 | 0.29 | |
| | | | | sex (male) | 0.39 | 1.37 | 0.29 | |
| | | | | timepoint FU:intervention prebiotic | -11.8 | 1.77 | -6.67 | **<0.001** |

*Abbreviations: FU - follow-up.*

#### 10. Spearman Correlation of neural response with microbiota and blood outcomes at first study visit

**Supplementary *Table 8: Spearman correlation analyses between the neural activations of specific clusters (left parahippocampus/amygdala and left parahippocampus) during retrieval at first study visit and fecal and serum SCFAs and gut microbiota.*** *Significant associations between clusters and SCFAs are shown, however, no association with* gut microbiota diversity, and or phylum was found, and only associations between clusters and specific genera and families are presented, for significant p-value. Microbiota associations did not survive FDR correction across included microbial families and genera. *FDR-correction was applied within outcome categories.*

| Neural activity of 2nd level (main) contrast at first study visit | marker (mean) | r value | p value | FDR p value |
|---|---|---|---|---|
| Left parahippocampus/amygdala | SCFA (mg) | 0.15 | 0.290 | 0.435 |
| | SCFA (serum, sum) | 0.01 | 0.962 | 0.962 |
| | SCFA (feces, sum) | 0.34 | **0.032** | 0.096 |
| | fecal Butyrate | 0.32 | **0.047** | 0.070 |
| | fecal Acetate | 0.27 | 0.094 | 0.094 |
| | fecal Propionate | 0.38 | **0.017** | **0.04995** |
| Left parahippocampus | SCFA (mg) | 0.38 | **0.005** | **0.014** |
| | SCFA (serum, sum) | 0.01 | 0.955 | 0.955 |
| | SCFA (feces, sum) | 0.18 | 0.275 | 0.412 |
| | fecal Butyrate | 0.14 | 0.387 | 0.387 |
| | fecal Acetate | 0.20 | 0.208 | 0.312 |
| | fecal Propionate | 0.21 | 0.203 | 0.312 |
| Left parahippocampus/amygdala | Christensenellaceae (R.7.group) | -0.32 | **0.017** | 0.756 |
| Left parahippocampus | Anaerofilum | -0.28 | **0.041** | 0.964 |

*Abbreviations: SCFA – short-chain fatty acid.*

#### 11. Spearman correlation of neural response with microbiota and blood outcomes

**Supplementary *Table 9: Spearman correlation analyses between the main neural activations during encoding and retrieval and fecal and blood markers.*** *FDR correction was applied within outcome categories. Only significant effects are shown here; the extended Table is shown in* Supplementary *Table 13. Hippocampus activity was extracted after second-level analyses (see "extr. Hippocampus act.") or an explicit mask (Tian et al., 2020) was used within second-level analyses (see hippocampus mask).*

| Neural activity of 2nd level (main) contrast | Fecal and blood markers (mean) | r value | p value | FDR p value |
|---|---|---|---|---|
| encoding food>art | SCFA (feces, sum) | -0.19 | **0.020** | 0.061 |
| hippocampus mask - encoding*wanting | SCFA (feces, sum) | -0.22 | **0.006** | **0.017** |
| encoding | serum Propionate | 0.21 | **0.035** | 0.141 |
| encoding*wanting | serum Lactate | 0.16 | **0.044** | 0.178 |
| hippocampus mask - retrieval | serum Lactate | -0.21 | **0.012** | **0.048** |
| encoding food>art | fecal Acetate | -0.23 | **0.004** | **0.011** |

| | | | | |
|---|---|---|---|---|
| hippocampus mask - encoding*wanting | fecal Butyrate | -0.27 | **0.001** | **0.002** |
| encoding | HCRP | -0.19 | **0.011** | 0.054 |
| retrieval | TSH | -0.17 | **0.030** | 0.074 |
| retrieval | IL6 | 0.17 | **0.026** | 0.074 |
| retrieval (food) | TSH | -0.16 | **0.035** | 0.173 |
| encoding*wanting | IL6 | 0.17 | **0.022** | 0.109 |
| retrieval*wanting | TSH | 0.21 | **0.006** | **0.028** |
| encoding | Creatinine | 0.19 | **0.010** | **0.039** |

*Abbreviations: HCRP* - High-Sensitivity C-Reactive Protein*, TSH* - Thyroid-Stimulating Hormone*, IL6* - Interleukin-6*.*

**Supplementary** ***Table 10: Spearman correlation analyses between the change of neural activations (baseline - post-prevention difference, delta) during encoding and retrieval (interaction) and the change of fecal and blood markers.*** *FDR correction was applied within outcome categories. Only significant effects of the prebiotic group compared to the placebo group are shown; the extended Table is shown in* Supplementary *Table 14. Hippocampus activity was extracted after second-level analyses (see "extr. Hippocampus act.") or an explicit mask (Tian et al., 2020) was used within second-level analyses (see hippocampus mask).*

| Group | Neural activity change (food> art, post-pre) of 2nd level (interaction; group*time) | Fecal and blood markers change (post-pre) | r value | p value | FDR p value |
|---|---|---|---|---|---|
| Prebiotic | retrieval (extr. hippocampus act.) | Evenness | -0.38 | **0.019** | 0.057 |
| Placebo | retrieval (extr. hippocampus act.) | Evenness | -0.03 | 0.868 | 0.868 |
| Prebiotic | retrieval | FM | -0.34 | **0.031** | 0.124 |
| Placebo | retrieval | FM | -0.10 | 0.550 | 0.638 |
| Prebiotic | encoding | Methane.metabolism..ko00680. | -0.34 | **0.024** | 0.122 |
| Prebiotic | encoding (cluster 1) | Methane.metabolism..ko00680. | -0.34 | **0.028** | 0.142 |
| Prebiotic | encoding (extr. hippocampus act.) | Flavonoid.biosynthesis..ko00941. | -0.31 | **0.046** | 0.229 |
| Placebo | encoding | Methane.metabolism..ko00680. | 0.19 | 0.216 | 0.328 |
| Placebo | encoding (cluster 1) | Methane.metabolism..ko00680. | 0.15 | 0.352 | 0.465 |
| Placebo | encoding (extr. hippocampus act.) | Flavonoid.biosynthesis..ko00941. | 0.18 | 0.257 | 0.570 |
| Prebiotic | hippocampus mask - encoding | Firmicutes | 0.42 | **0.006** | **0.012** |
| Prebiotic | encoding (extr. hippocampus act.) | Firmicutes | 0.38 | **0.013** | **0.025** |
| Placebo | hippocampus mask - encoding | Firmicutes | 0.05 | 0.749 | 0.749 |
| Placebo | encoding (extr. hippocampus act.) | Firmicutes | 0.08 | 0.586 | 0.677 |
| Prebiotic | retrieval | Lachnospiraceae.NK4A136.group | 0.32 | **0.044** | 0.133 |
| Prebiotic | retrieval (extr. hippocampus act.) | UCG.003 | 0.38 | **0.018** | 0.053 |
| Prebiotic | retrieval (old) (extr. hippocampus act.) | UCG.003 | 0.35 | **0.031** | 0.092 |
| Placebo | retrieval | Lachnospiraceae.NK4A136.group | -0.05 | 0.786 | 0.786 |
| Placebo | encoding (extr. hippocampus act.) | Lachnospiraceae.NK4A136.group | 0.15 | 0.343 | 0.353 |
| Placebo | encoding (extr. hippocampus act.) | Lachnospiraceae.FCS020.group | 0.14 | 0.353 | 0.353 |
| Placebo | encoding (extr. hippocampus act.) | UCG.003 | 0.15 | 0.344 | 0.353 |
| Placebo | retrieval (extr. hippocampus act.) | Lachnospiraceae.NK4A136.group | -0.03 | 0.842 | 0.842 |
| Placebo | retrieval (extr. hippocampus act.) | Lachnospiraceae.FCS020.group | 0.26 | 0.120 | 0.180 |
| Placebo | retrieval (extr. hippocampus act.) | UCG.003 | -0.31 | 0.060 | 0.179 |
| Placebo | retrieval (old) (extr. hippocampus act.) | Lachnospiraceae.NK4A136.group | -0.16 | 0.342 | 0.910 |
| Placebo | retrieval (old) (extr. hippocampus act.) | Lachnospiraceae.FCS020.group | -0.02 | 0.910 | 0.910 |

| | | | | |
|---|---|---|---|---|
| Placebo retrieval (old) (extr. hippocampus act.) | UCG.003 | -0.03 | 0.842 | 0.910 |
| Prebiotic encoding | Enterococcus | -0.31 | **0.045** | 0.224 |
| Prebiotic encoding | Erysipelatoclostridium | -0.42 | **0.006** | 0.058 |
| Prebiotic retrieval (old) | g_Unknown | 0.33 | **0.040** | 0.397 |
| Prebiotic encoding (cluster 1) | Erysipelatoclostridium | -0.42 | **0.006** | 0.063 |
| Prebiotic retrieval (old) (cluster 1) | g_Unknown | 0.33 | **0.041** | 0.413 |
| Prebiotic retrieval (old) (extr. hippocampus act.) | Oscillospira | -0.34 | **0.035** | 0.180 |
| Prebiotic retrieval (old) (extr. hippocampus act.) | Peptostreptococcus | 0.34 | **0.036** | 0.180 |
| Placebo encoding | Enterococcus | 0.08 | 0.600 | 0.800 |
| Placebo encoding | Erysipelatoclostridium | 0.23 | 0.134 | 0.800 |
| Placebo retrieval (old) | g_Unknown | -0.13 | 0.439 | 0.924 |
| Placebo encoding (cluster 1) | Erysipelatoclostridium | 0.23 | 0.128 | 0.868 |
| Placebo retrieval (old) (cluster 1) | g_Unknown | -0.15 | 0.381 | 0.854 |
| Placebo retrieval (old) (extr. hippocampus act.) | Oscillospira | 0.03 | 0.865 | 0.933 |
| Placebo retrieval (old) (extr. hippocampus act.) | Peptostreptococcus | 0.15 | 0.390 | 0.747 |

*Abbreviations: FM – fat mass.*

#### 12. Memory encoding and retrieval of different stimulus categories

**Neural correlates of calorie-biased food memory**

We could not confirm that food memory encoding and retrieval is biased by caloric intake (H5_m) and neither that memory encoding can potentially be modulated by wanting of different caloric stimuli (H9_e).

Post-hoc exploratory analyses revealed that the encoding of food images which contain lower caloric contents (*cal 1 quantile*, see Medawar et al., 2024) is reduced after prebiotic intervention compared to placebo (*beta=-1.19, SE=0.46, T=-2.6,* $x^2$ *p=0.009; Figure S4A, Table S6). Furthermore, overall the encoding of the lowest an*d the highest caloric quantile was significantly different (*p=0.0313, F(78)=4.8105*).

Moreover, the memory accuracy during retrieval of fruit stimuli (*Food Type 2*) was reduced after prebiotic intervention (*beta=-0.57, SE=1.05, T=-0.54, x2 p=0.003;* Figure S4B). During retrieval, this decline in the memory accuracy of fruit stimuli after the intervention was inversely associated with a higher reduction of neural activations in (see Figure 3B; *rho=-0.47, pFDR=0.0024;* Figure S4C).

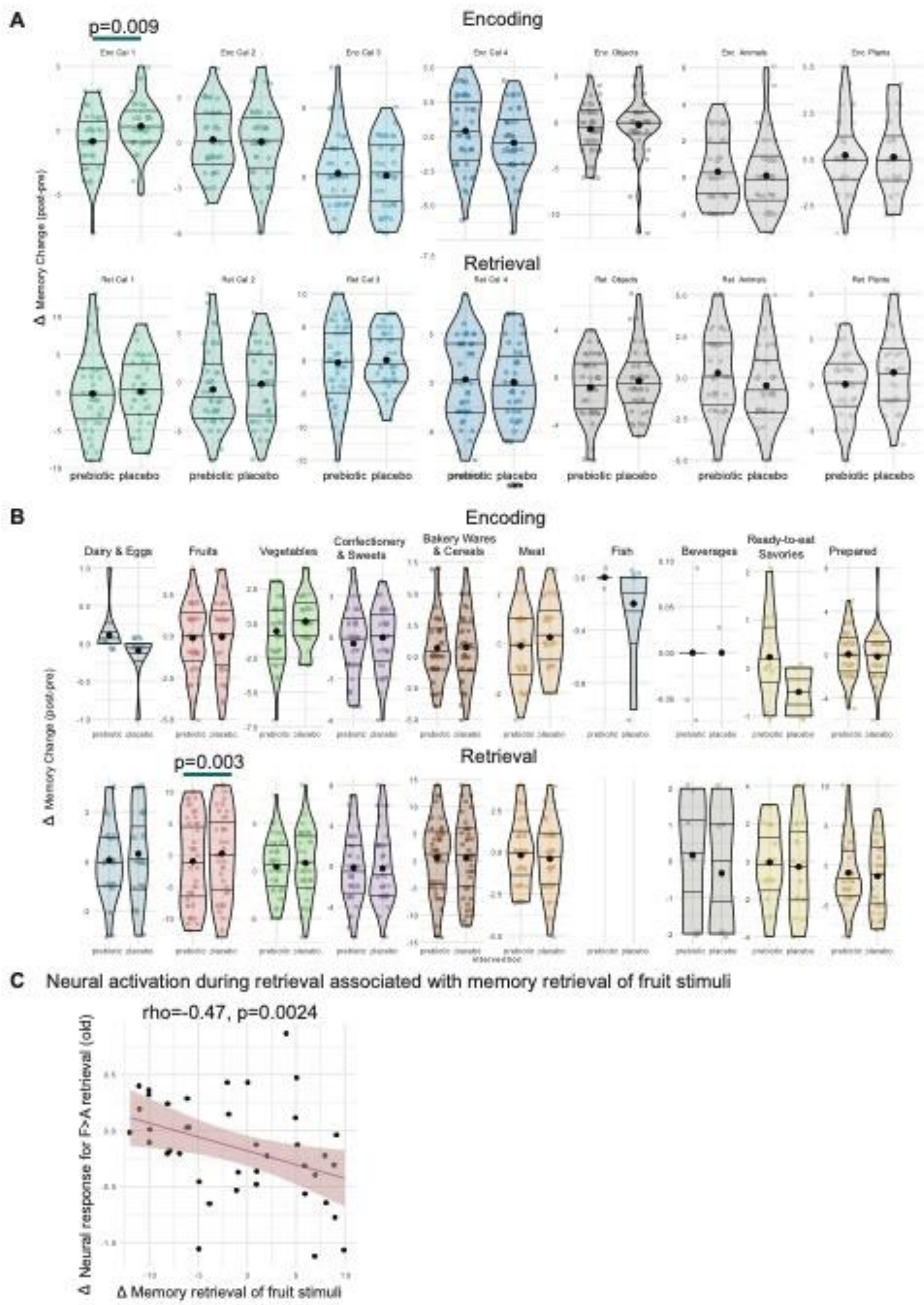

***Supplementary Figure 4: Change of successful memory encoding and retrieval within diet categories and food types after intervention.*** *Memory accuracy of (A) food and art stimuli categories and (B) food stimuli separated into nutritional food types. (C) Scatterplot of Spearman correlation between the change of memory retrieval of food stimuli type 2 (fruits) and the neural activation during retrieval of old images.*

#### 13. Supplementary Discussion

Prebiotic intervention showed a stronger association with neural responses beyond the dmPFC, ACC, and OFC during the correct identification of old food versus art stimuli retrieval. Specifically, the thalamus integrates sensory information and supports limbic system relay. The OFC, in turn, orchestrates motivated food intake by combining sensory and reward-related inputs. Thus, decreased thalamic and OFC responses may reflect attenuated homeostatic–hedonic integration during food retrieval. Similarly, reduced hypothalamic engagement supports the interpretation that prebiotics modulated homeostatic signaling during retrieval. Other frontal regions, including the dmPFC and ACC, are central for reward pathway modulation and interact with dopaminergic input from the VTA. The caudate and hippocampus further contribute to stimulus–reward learning and memory-guided behavior. In food cue tasks, caudate activation is often linked to reward anticipation and wanting[1], suggesting that reduced caudate activity may reflect diminished habit-related or reinforcement-driven processes. The hippocampus, beyond its established role in memory, also links hippocampal-dependent memory processes of learned food stimuli with internal states of hunger, satiety, and appetitive control[2,3]. Structural evidence further underscores its importance, as reduced hippocampal grey matter volume has been observed in individuals with obesity[2]. Furthermore, we previously reported in a large longitudinal cohort that lower diet quality and abdominal obesity in midlife were associated with reduced hippocampal functional connectivity and altered white matter integrity[4].

#### 14. Details on Model Designs

In the first-level step, we used four predictors (placebo_BL, placebo_FU, prebiotic_BL, prebiotic_FU) and eight design matrices. Memory accuracy ('correct') was estimated for each predictor using the correct identification of target images during encoding and retrieval and the correct rejection of novels and lures images during retrieval.

- Neural activations related to memory accuracy were estimated using design matrix encoding and retrieval: memory correct [0.5 0 0.5 0], food memory correct [1 0 0 0], art memory correct [0 0 1 0], food > art [1 0 -1 0], art > food [-1 0 1 0].
- Neural activations related to wanting modulated memory accuracy were estimated using design matrix encoding_wanting and retrieval_wanting: memory*wanting slope [0 0.5 0 0.5], food memory*wanting slope [0 1 0 0], art memory*wanting slope [0 0 0 1].
- Neural activations related to caloric bias (kcal modulation) memory accuracy were estimated using design matrix encoding_kcal and retrieval_kcal: memory*kcal [0 0 1 0], kcal [0 1 0 0]. For more specific contrasts on the nutrient level, to test the intervention effect of memory accuracy modulated by nutrient value, we will use GLMs with additional interaction regressors (1 in design matrix): memory + kcal_100g + kcal_100g*memory as regressors

- Additionally, neural activations related to kcal and wanting modulated memory accuracy were estimated using design matrix encoding_wantingkcal: memory*wanting slope*kcal [0 0 1 0], kcal*wanting slope [0 0.5 0 0.5].
- Moreover, we included models during retrieval including only correct identification of target ('old') stimuli using design matrix retrieval_old: memory old stimuli correct [0.5 0 0.5 0], food old stimuli memory correct [1 0 0 0], art old stimuli memory correct [0 0 1 0], food > art [1 0 -1 0], art > food [-1 0 1 0]; and retrieval_old_wanting: memory old stimuli *wanting slope [0 0.5 0 0.5], food memory old stimuli *wanting slope [0 1 0 0], art memory old stimuli *wanting slope [0 0 0 1].
- Moreover, to test H6_m (in preregistration) we repeated 1st level SPM models encoding and retrieval using only baseline (BL) predictors for the first study visit (ses-01).

Orthogonalizations in SPM were set to 1 in case of one parametric modulator per condition (models encoding, retrieval) and in case of more than one parametric modulator, orthogonalisation was set to 0 (models encoding_wanting, retrieval_wanting, encoding_kcal and retrieval_kcal, encoding_wantingkcal).

During 2$^{nd}$ level analyses, we used the explicit mask consisting of a priori selected ROIs and the reward network mask (see ROI selection). We repeated the following models using an explicit hippocampus mask consisting of head, body, and tail hippocampus subregions [5] : encoding, retrieval, wanting, retrieval_wanting, encoding_kcal, retrieval_kcal

**Main effects to test Hypotheses to test main hypotheses H1_m to H5_m and explorative hypotheses H1_e, H2_e, and H6_e:** Food viewing elicits different BOLD responses compared to art viewing. We will report overall main effects on the 2nd level (T-test with the following contrast matrix to assess the mean of all conditions and timepoints). We note that for encoding and retrieval models at first study visit (ses-01), we only test main effects.

Contrast Matrix: [0.25 0.25 0.25 0.25]

**Intervention effects to test the explorative hypotheses H3_e to H9_e:** Food compared to art memory accuracy elicits different BOLD responses when comparing pre/post-intervention. There is an interaction of timepoint and condition on the first-level contrasts.

Contrast Matrix: [-1 1 1 -1]

#### 15. tSNR of ROIs

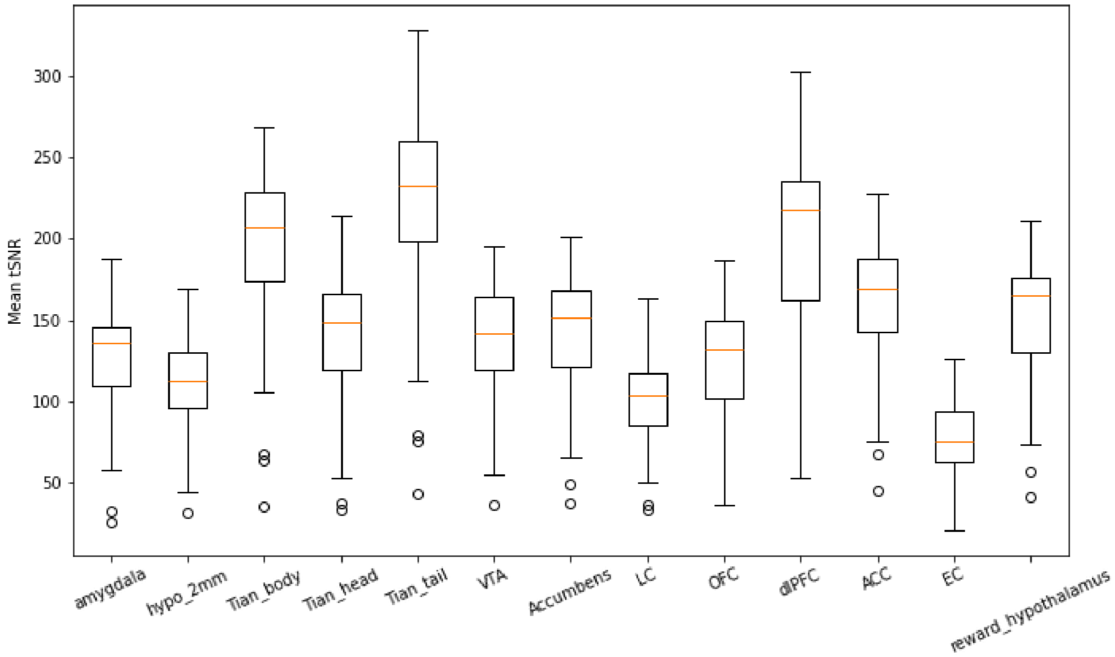

***Figure S5: Region-specific mean tSNR during memory task-fRMI across all four sessions and time course.*** *Hypo – hypothalamus, Tian_body, head, tail – hippocampus segmented body head and tail by Tian et al., 2020, VTA - ventral tegmentum area, LC – locus coeruleus, OFC – orbitofrontal cortex, dlPFC – dorsolateral prefrontal cortex, ACC – anterior cingulate cortex, EC – entorhinal cortex, reward hypothalamus – mask by Medawar et al., 2024 including hypothalamus and reward areas.*

#### References

1. Kanoski SE, Boutelle KN. Food cue reactivity: Neurobiological and behavioral underpinnings. *Rev Endocr Metab Disord*. 2022;23(4):683-696. doi:10.1007/s11154-022-09724-x

2. Barbosa DAN, Gattas S, Salgado JS, et al. An orexigenic subnetwork within the human hippocampus. *Nature*. 2023;621(7978):381-388. doi:10.1038/s41586-023-06459-w

3. Stevenson RJ, Francis HM, Attuquayefio T, et al. Hippocampal-dependent appetitive control is impaired by experimental exposure to a Western-style diet. *Royal Society Open Science*. 2020;7(2):191338. doi:10.1098/rsos.191338

4. Jensen DEA, Ebmeier KP, Akbaraly T, et al. Association of Diet and Waist-to-Hip Ratio With Brain Connectivity and Memory in Aging. *JAMA Netw Open*. 2025;8(3):e250171. doi:10.1001/jamanetworkopen.2025.0171

5. Tian Y, Margulies DS, Breakspear M, Zalesky A. Topographic organization of the human subcortex unveiled with functional connectivity gradients. *Nat Neurosci*. 2020;23(11):11. doi:10.1038/s41593-020-00711-6
